# Advancing Breast Cancer Detection: A Comprehensive Evaluation of Machine Learning Models on Mammogram Imaging

**DOI:** 10.1101/2025.10.08.25337620

**Authors:** Reshad Al Muttaki, Sadia Afrin, Alvi Ibn Amzad Anil, Mehedi Hasan Shawon

## Abstract

Breast cancer, which is among the top causes of cancer-related deaths in women worldwide, demonstrates the importance of effective and rapid diagnostic tools, especially in early diagnosis, to enhance the survival level. Although machine learning (ML) advances have had an increasing number of medical imaging applications, limitations of diversity and applicability of datasets, the interpretation and efficiency of models remain a challenge to clinical use. The paper assesses eight of the most popular ML models, such as Convolutional Neural Network (CNN), Kolmogorov-Arnold Network (KAN), k-Nearest Neighbors, Support Vector Machine, XGBoost, Random Forest, Naive Bayes, and a Hybrid model based on the Mammogram Mastery dataset of Iraq-Sulaymaniyah, which consists of 745 original and 9,685 augmented mammogram images. The hybrid model has the best accuracy (0.9667) and F1 Score (0.9444), and the KAN model has the best ROC AUC (0.9760) and Log Loss (0.1421), meaning they are best in terms of discriminative power and proper calibration. Random Forest, which has the lowest false negatives (3) when compared with Fast Multinomial and Fast Text, became most secure in clinical screening since it struck a balance between sensitivity and computing efficiency. The two practical challenges, though, are the slow inference time of the KAN model (0.323 seconds) and the expensive training cost (1009.10 seconds) of the Hybrid model. These insights explain that the Hybrid and KAN models are promising means of improving the accuracy of the diagnostics, and Random Forest can serve as a practically representative tool for reducing the number of missed diagnoses. The context of future research needs to address multi-dataset validation from multiple institutions, speed optimization of inference, multi-classification, and improved interpretability that will be used in clinically integrative settings. By addressing these gaps, ML-based diagnostics have the potential to increase the rate of breast cancer diagnosis, minimizing diagnostic errors and improving patient outcomes in various clinical contexts, which can facilitate the scaling of screening services available across the world.

## 1. Introduction

Breast cancer is currently one of the most concerning health issues on a global scale, being the most prevalent cancer in women and the cause of the highest number of cancer deaths worldwide. According to World Health Organization (WHO), it is reported that around 2.3 million new cases of breast cancer were diagnosed in 2022, with 670,000 individuals dying around the world due to the disease, illustrating the high burden it causes [1]. In the US in 2024, it is estimated that over 310,720 women are to be diagnosed with invasive breast cancer, with another 56,500 instances of in situ breast cancer, further emphasizing the importance of effective screening and diagnostic measures [2]. The mortality rate can be reduced through an early diagnosis, and early-stage breast cancer (stages I and II) has a survival rate of more than 90 percent at 5 years, as opposed to a little bit more than 30 percent in the case of advanced-stage (stage IV) cancer [3]. Mammography, which has been the pillar behind breast cancer screening, has greatly diminished the number of mortalities since it can detect the tumor before it becomes palpable. Yet, the radiologist fatigue, inter-observer variability, and difficulty in the detection of subtle lesions, especially those with dense breast tissue in women, are some of its shortcomings [4]. This is made difficult in resource-poor locations where there may not be access to skilled radiologists and high-tech imaging machines. Machine learning (ML) provides a revolutionary solution to this part and replenishes image analysis automation, which improves the accuracy of diagnosis and allows widespread screening programs to address access imbalances in healthcare.

This research is motivated by the dire demand to generate sound, precise, computationally tractable ML models that aid in early identification of the breach of breast cancer when it is still at its most treatable levels. Mammogram interpretation is mammography in general is entirely subjective, and literature suggests that radiologists have a 10-30% likelihood of missing cancer because it is not perceptually caught due to fatigue [5]. False positive indications also tend to create unwarranted biopsies and panic in patients. One of the major revolutions made by ML and, in particular, deep learning is the application in the field of medical imaging, where the feature extraction and pattern detection that it facilitated can be automated and outperform human performance in terms of consistency and speed. This work aims to find solutions to the most urgent issues related to breast cancer detection, employing better sensitivity to early-stage lesions, lower false positivity, and generalizability among various patients and imaging settings using sophisticated ML models. Besides, there are new ML frameworks, like Kolmogorov-Arnold Networks (KANs), that are also on the agenda to be discussed during the research. Another aspect of the study is the implementation of hybrid methods combining deep learning with classical ML approaches, promoting high diagnosis accuracy and clinical explanations. Fig 1 shows how a typical breast cancer detection system using ML looks like.

**Fig 1.**
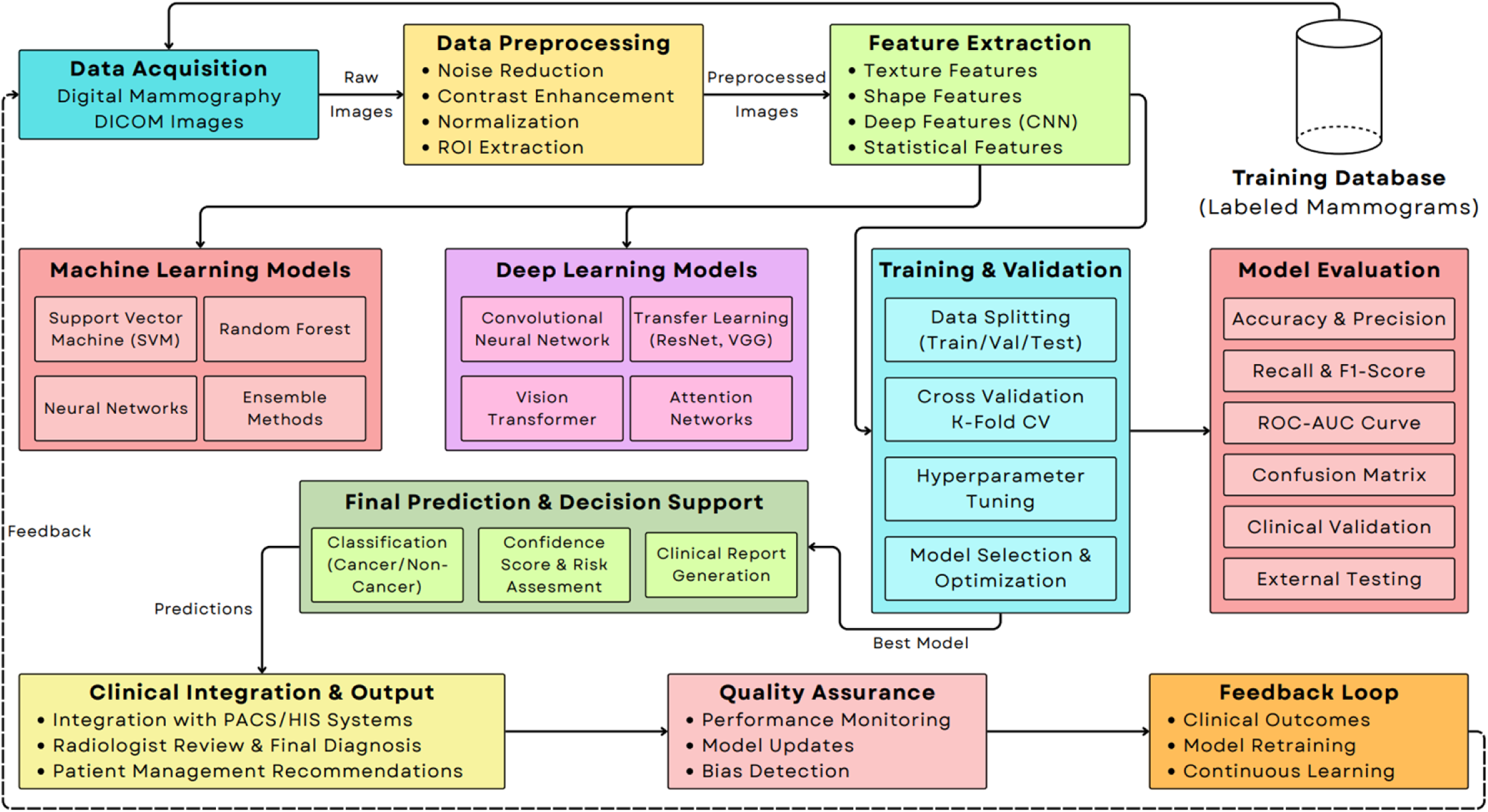
A Typical Breast Cancer Detection System using Machine Learning.

The power of CNNs to handle simple and complex datasets, such as mammograms, lies in their ability to extract hierarchical spatial information through convolution. Due to this, CNNs are becoming the basis of medical Image analysis. InceptionV3, VGG16, DenseNet121, and ResNet50 are models that yield superior results when detecting significant mammographic components, including microcalcifications, masses, and architectural changes, which can be major signs of malignancy [6–8]. Such deep architectures perform well using multiple layers to learn detailed patterns and are thus suitable for the case of subtle abnormalities detection in high-resolution images. Additionally, the robustness is boosted due to ensemble models that combine the predictions of multiple classifiers (e.g., Random Forest, Support Vector Machines, or multiple CNNs), reducing the impact of the overfitting issue and strengthening generalization to different datasets [7,9,10]. A new type of model is Vision Transformers (ViTs), which exploit the power of attention to model global dependencies on images, providing a complement to CNNs since they can model long-range connections, which might be important to the analysis of a family of mammograms holistically [11]. The models are emerging in cases where the global context, i.e., breast tissue asymmetry, contributes to the diagnosis. Also, less popular traditional ML models such as Random Forest, Logistic Regression, or Support Vector Machines still exist and are applicable in such settings as hybrid frameworks to augment deep learning feature extractors [10,12,13].

Kolmogorov-Arnold representation is a breakthrough in ML based on the Kolmogorov-Arnold theorem of representation, which mathematically proves the fact that any continuous multivariate function can be realized as a composition of univariate functions. In contrast with traditional neural networks that use fixed penalty functions (e.g., ReLU) and linear weight matrices, KANs have learnable univariate functions that allow them to capture more natural and high complexity non-linear relations in a more flexible and interpretable manner. Such flexibility is specifically useful in the field of medical imaging, where the variability observed in most datasets may be considerable because of different imaging devices, patient inspection, and ailment presentation. KANs have also been used in breast cancer detection, with some work being done to add them to Hybrid architectures, e.g., the U-KAN model that adds KAN blocks to a U-Net framework and has shown greater visibility and increased segmentation accuracy when detecting lesions [11]. Even though KANs remain in early development, they have the potential to show promise in allowing mammography tests to be less complex and still perform as well or better, an opportunity that could make them useful in achieving improved precision of a diagnostic test.

Hybrid models are promising solutions within breast cancer detection, where the information extracted by various deep learning or machine learning models (e.g., CNN, ResNet, DenseNet, etc.) is paired with a traditional ML classifier (e.g., SVM, Random Forest, etc.) or other advanced neural networks. These models combine the image pattern learning capabilities of deep learning with the interpretable predictions of traditional classifiers, through the automated feature learning property of deep learning to capture complex image patterns. Researchers have demonstrated that the methods of hybrid solutions, e.g., combining ResNet50 and SVM or DenseNet121 with the ensemble classifier, may be better at handling issues such as overfitting, dataset fluctuations, and poor interpretability as compared to isolated methods [9,14,15]. As an example, hybrid models have the potential to reduce heavy computation requirements of deep learning, incorporating deep-trained CNNs as feature analyzers and lightweight classifiers, rendering them practical enough to be deployed in resource-limited situations in clinical practice [13]. What they do not fully emphasize, however, is that hyperparameter tuning, such as Genetic Algorithms (GA) or Particle Swarm Optimization (PSO), can further optimize the results of the hybrid model, since it was proven in studies that improved Random Forest and CNN-based frameworks [16,17]. These models are especially suitable in mammogram analyses where raw data from various sources and the explainability of the predictions are essential aspects before the clinical use of the models.

### Main Contributions of This Research

- Creation of a new mosaic model that entails a CNN Feature Extractor with a 1024-D feature vector and a weighted combination of CNN, Random Forest, and XGBoost classifiers, which attain higher accuracy and F1 score.
- A complete analysis of the results of eight ML models, both classical classifiers (e.g., KNN, Naive Bayes) and more recent DL frameworks (e.g., KAN), with a standard performance matrix.
- Recognition of potential areas of future research (including the insufficient data variety and no integration of multi-modal data) and their resolutions.

This paper presents a comprehensive analysis of several ML models on breast cancer identification with the Mammogram Mastery dataset, consisting of 745 original and 9,685 augmented mammogram images obtained in Northern Iraq, Sulaymaniyah [18]. The comparison of CNNs, ensemble models, ViTs, KANs, and hybrid models should thus provide an insight into the most appropriate methods to enhance the accuracy of diagnoses and how the research in these fields can be improved (namely through the increased diversity of datasets, interpretability, and cross-modal applicability). The consideration of KANs and hybrid models is a new addition, which has potential as it appears to eliminate the flaws of traditional models. In our results, we aim to identify prospective diagnostic tools that are dependable, continuous, and explainable in the pursuit of finding a solution to enhance clinical decision-making, reduce diagnostic errors, and improve patient outcomes in breast cancer management. Moreover, harnessing an example of a unique regional view on the dataset, this study will help to fill the worldwide gap in the research on breast cancer prevalence and contribute to the medical education of a professional population.

## 2. Related Works

Machine learning (ML) and deep learning (DL) transform the possibility of detecting breast cancer, solving the current problem of the lack of adequate tools that help radiologists analyze complex medical images due to their large scale and precision mismatch. The literature is diverse, both in terms of the methodology coverage, including classical ML classifiers to complex DL models, and the imaging modalities, as mammography, histopathology, ultrasound, thermography, etc. The various studies are aimed at making early detection, better lesion separation, and also correct diagnosis between benign and malignant tumors, and issues such as dataset variability, interpretability of the models, and clinical applicability are addressed. The subsection summarizes the main insights of recent research studies on their methodology, contributions, and limitations in order to draw a complete picture within the area.

Fofnvik et al. used a 3D ResNet (MedicalNet: 18 layers) with an attention module to determine serial digital breast tomosynthesis in a prospective cohort of breast cancer patients following neoadjuvant chemotherapy [6]. Their model had the potential to predict treatment response but had limitations of a small cohort and single-institution data, and had to be validated in a wider setting. Balkenende et al. have extensively reviewed DL uses in breast cancer imaging, including convolutional neural networks (CNN) such as GoogLeNet, YOLO, DenseNet, Faster R-CNN, U-Net, Inception-v3, and AlexNet, Generative Adversarial Networks (GAN), and Deep Belief Networks (DBN) [14]. Their effort highlights how versatile these models can be with ultrasound and MRI, but puts a major focus on the necessity of widespread clinical trials, especially in the context of the nuclear medical field.

Huang et al. designed a U-Net-like network whose convolutional layers were based on MobileNetV2 to improve the performance of detecting various images in complex histopathology images using a novel cross-staining workflow (CDACS) [19]. Alotaibi et al. concentrated on ultrasound imaging, where VGG19 and EfficientNet V2 are used to do the classification, and Mask-RCNN is used to auto-crop images, but also to segment [20]. The work by Solorzano et al. deployed ten individual instances of an object detection model, InceptionV3, to detect invasive breast cancer in histopathology images, but with only a small sample size (587), as whole-slide images (WSIs) [7]. Qasrawi et al. made the suggestion of a hybrid ensemble model using the DenseNet121 neural network in feature extraction and ensemble deep Random Vector-Functional Link Neural Network (edRVFL), which includes YOLOv5 and MedSAM in lesion detection/segmentation [9]. The model they fabricated had better clinical performance, yet they had difficulty with the variability of imaging equipment.

Pertuz et al. have tested the four AI systems End2End, DMV-CNN, GMIC, and GLAM on Grad-CAM with the assistance of saliency analysis, which showed they still exhibit interpretability difficulties [21]. Comparing 6 classical ML models, including RF, DT, and SVC, Khalid et al. mentioned that most of the cases incurred a lot of computational expenses [12]. Zhu et al. proposed a U-KAN framework that combines Kolmogorov-Arnold Networks (KANs) and U-Net to perform lesion segmentation better and support interpretability [11]. Ravi Kumar et al. assessed nine traditional ML classifiers, including RF and XGBoost, on tabular data, recommending the direction of their possible application in the future on images [10].

Essa et al. used an approach in which the values of biomarkers are converted into images to overcome the problem of biomarker variability by using a multi-stage pipeline with ResNet50 as the classifier [15]. Rabiei et al. used RF, Multi-layer Perceptron (MLP), and Gradient Boosting Trees (GBT) and Genetic Algorithm (GA) optimization, but were constrained to one database [16]. Veerlapalli and Dutta offered a BCDGAN model that combines GANs with a mixture of CNNs and VGG16, InceptionResNetV2 hybrid classifier, and is tested over one dataset [8]. Twum et al. have used transfer learning by utilizing MobileNetV2, InceptionV3, ResNet50, and classifiers such as LightGBM, and gave warnings of overfitting [13]. Sinjanka et al. incorporated clinical, genetic, and lifestyle data with RF and CNNs and were restricted by the sample of a small region [22].

Naji et al. involved five ML classifiers in comparison with the Wisconsin Breast Cancer Diagnostic dataset, which demanded wider validation [23]. Singh et al. combined features of VGG16 and hand-designed CNN, and identified by an ensemble of SVM and Decision Trees, which made high accuracy [24]. Lilhore et al. proclaimed a combination of CNN, Bi-LSTM, and EfficientNet-B0 model to accomplish high accuracy classification [25]. Zhu et al. integrated SHAP, Recursive Feature Elimination (RFE), and RF with Particle Swarm Optimization (PSO) to detect early [17]. Shen et al. transformed ResNet50 and VGG16 into classifiers that operate on the entire image, with the disadvantage of image shrinking [26].

Sechopoulos et al. assessed the state-of-the-art of AI in mammography, comparing Deep Learning (DL) models such as CNN and YOLO to conventional and hand-crafted feature systems and noted the requirement of further large-scale prospective trials and the difficulty in including temporal comparisons to the previous examinations [27]. Xing et al. applied a pre-trained ResNet34 and Logistic Regression on Contrast-Enhanced Spectral Mammography (CESM) to predict the neoadjuvant chemotherapy response, and acknowledged the limitations of the study of using a small, single-center, retrospective dataset [28]. Al Mansour et al. used transfer learning on NasNetMobile, EfficientNet-b0, and MobileNetV2 on the digital mammograms, which suggests the necessity of the elevated accuracy and explainability of the models used in clinical practice [29]. Chitta et al. proposed a hybrid model consisting of a schema-specific EfficientNetV2 and a Vision Transformer (ViT) to enhance the accuracy of diagnosing a biopsy using a histopathological image, which, based on the results, should be evaluated using larger datasets [30]. In order to deal with the low sensitivity of mammography in dense tissue, Gupta et al. developed a diagnostic framework based on Disentangled Variational Autoencoder (D-VAE) and Fully Elman Neural Network (FENN) to combine the features of mammogram and Digital Breast Tomosynthesis (DBT) [31]. Talukdar et al. trained a hybrid model combining the characteristics of Xception and EfficientNet-B5, following the enhancement of images using an autoencoder, and admitted that the use of a relatively small database constrained segmentation performance [32]. Sansone et al. have used the classical ML models such as SVM and Random Forest in estimating the amount of breast density using radiomic features of mammograms, admitting the preliminary nature of the study since the dataset was small [33]. In an overall analysis, Hanis et al. revealed that even the models such as Neural Networks, SVM, etc. are widespread, their application in the real context of the clinical process is concealed with the problems of privacy, trust, and unambiguous clinical roles [34]. Different DL architectures, such as CNNs and GANs, were surveyed by Wang, who stressed the lack of large and high-quality datasets and data annotation and interpretation difficulties [35]. Alshammari et al. in their pilot study have compared classical ML classifiers such as KNN and SVM on a small and local dataset and advised further experiments using more sophisticated individual and hybrid models [36].

Table 1 summarizes key studies, their ML models, imaging modalities, key findings, limitations, and citations, providing a comprehensive overview of the field.

**Table 1.** Overview of ML Models in Breast Cancer Detection.

| Study | ML Models Used | Imaging Modality | Key Findings | Limitations | Citation |
| --- | --- | --- | --- | --- | --- |
| Förnvik et al. | 3D ResNet<br>(MedicalNet) | Tomosynthesis | Predicts treatment response | Small cohort, single institution | [6] |
| Balkenende et al. | CNNs, GANs, DBNs | Mammography, Ultrasound, MRI | Versatile across modalities | Needs large-scale trials | [14] |
| Huang et al. | MobileNetV2 U-Net | Histopathology | CDACS enhances accuracy | Relies on nuclei information | [19] |
| Alotaibi et al. | VGG19, EfficientNet V2, Mask-RCNN | Ultrasound | Preprocessing improves performance | Manual cropping needed | [20] |
| Solorzano et al. | InceptionV3 Ensemble | Histopathology | Ensemble improves accuracy | Small sample size | [7] |
| Qasrawi et al. | DenseNet121, edRVFL, YOLOv5, MedSAM | Mammography | Superior clinical performance | Device variability | [9] |
| Pertuz et al. | End2End, DMV-CNN, GMIC, GLAM | Mammography | Saliency analysis insights | Interpretability challenges | [21] |
| Khalid et al. | RF, DT, SVC, KNN, LR | Tabular Data | Effective for tabular data | High computational demands | [12] |
| Zhu et al. | U-KAN, ViT | Mammography | Improved segmentation accuracy | Early-stage KAN adoption | [11] |
| Ravi Kumar et al. | RF, XGBoost, GBDT | Tabular Data | High accuracy on tabular data | Needs image-based studies | [10] |
| Essa et al. | ResNet50, SVM, Fuzzy NN | Biomarker Images | Novel feature engineering | Biomarker variability | [15] |
| Rabiei et al. | RF, MLP, GBT, GA | Tabular Data | GA optimization improves performance | Single database | [16] |
| Veerlapalli and Dutta | BCDGAN, InceptionResNetV2, VGG16 | Thermography | Enhanced feature extraction | Single dataset validation | [8] |
| Twum et al. | MobileNetV2, ResNet50, LightGBM | Ultrasound | Transfer learning effective | Risk of overfitting | [13] |
| Sinjanka et al. | RF, CNN | Multi-modal Data | Integrates diverse data | Small regional sample | [22] |
| Naji et al. | SVM, RF, KNN, DT | Tabular Data | Comparative analysis | Single database | [23] |
| Singh et al. | VGG16, Custom CNN, SVM | Mammography | Feature fusion improves accuracy | Computational complexity | [24] |
| Lilhore et al. | CNN, Bi-LSTM, EfficientNet-B0 | Mammography | High-accuracy classification | Real-time challenges | [25] |
| Zhu et al. | RF, SHAP, RFE, PSO | Tabular Data | Enhanced feature selection | High computational cost | [17] |
| Shen et al. | ResNet50, VGG16 | Mammography | Whole-image classification | Image downsizing issues | [26] |
| Sechopoulos et al. | DL, CNNs, YOLO | Mammography | DL models are state-of-the-art | Need for prospective trials, temporal comparison, medico-legal clarity | [27] |
| Xing et al. | ResNet34, Logistic Regression | CESM | DL features predict NAC response | Single-center, small sample, retrospective | [28] |
| Al Mansour et al. | NasNetMobile, EfficientNet-b0, MobileNetV2 | Mammography | Transfer learning shows promise | Lack of high accuracy, explainability, and diverse datasets | [29] |
| Chitta et al. | EfficientNetV2, Vision Transformer (ViT) | Histopathology | Hybrid model enhances diagnosis | Needs validation on larger, diverse datasets | [30] |
| Gupta et al. | D-VAE, FENN, Dg-CCA | Mammography, DBT | Fuses features from multiple modalities | High computational burden, need for real-time deployment | [31] |
| Talukdar et al. | Xception, EfficientNet-B5, U-Net | Mammography | Feature fusion improves classification | Suboptimal segmentation due to limited dataset | [32] |
| Sansone et al. | SVM, LDA, ANN, DT, RF | Mammography | Quantifies breast density objectively | Limited single-institution dataset | [33] |
| Hanis et al. | Neural Networks, SVM, DL models | Mammography | Meta-analysis of diagnostic accuracy | Publication bias, reliance on public datasets, lack of clinical role definition | [34] |
| Wang | CNNs, RNNs, GANs, Autoencoders | Mammography | Review of DL architectures | Scarcity of large, high-quality datasets, lack of interpretability | [35] |
| Alshammari et al. | DT, KNN, SVM, NB, DA | Mammography | Pilot study comparing classic ML models | Small local dataset (42 instances) | [36] |

### 2.1 Commonly Used Machine Learning Models

The literature in question shows a rather heterogeneous class of models of ML most frequently used in breast cancer detection, which proves how intricate medical imaging tasks can be. Convolutional Neural Networks (CNNs) are prevalent because they produce hierarchical spatial features of high-resolution images such as mammograms. ResNet50, because of the residual connections, overcomes vanishing gradient problems, which allows deep architectures to learn more complex patterns, like microcalcifications and masses [6,13,15]. The simple and deep architecture of VGG16 also makes it a good candidate for transfer learning that takes advantage of the pre-trained weights to adjust to medical imaging-related applications [8,20,26]. InceptionV3 uses the multi-scale feature processing in processing various sizes of lesions, enhancing the detection of different abnormalities [7]. The dense connection guarantees the reuse of features (which is not the case with other well-known models), making DenseNet121 perform better on complex datasets [9]. MobileNetV2 and EfficientNet V2 are lightweight models that trade image quality differentially, providing an acceptable accuracy within computational resources, a unique attribute to clinical environments with compromised capabilities [13,19,20].

ViTs have become a strong alternative where attention structures are used to capture long-range dependencies and address global semantics, such as the asymmetry of the breast tissues, which is essential in comprehensive breast tissue analysis in the mammograms [11]. Conventional ML models, such as Random Forest (RF) or Support Vector Machines (SVM), Logistic Regression (LR), k-Nearest Neighbors (KNN), and Decision Trees, are still effective on tabular data or in the case of a hybrid system in which they are used together with deep learning feature extractors [10,12,22,23]. Ensemble models lead to robustness through the use of multiple classifiers. This ensemble of ten InceptionV3 models of Solorzano et al. performed a majority vote to enhance the accuracy [7], whereas Qasrawi et al. combined DenseNet121 and edRVFL to achieve a higher rate of performance [9]. There is an upward trend toward hybridizing CNN feature extractors with classical classifiers. Twum et al. combined ResNet50 and VGG16 with LightGBM and XGBoost [13], or fused VGG16 with custom CNN features, SVM, altogether with Decision Trees [24], fused ResNet50 and custom CNN features with LightGBM and XGBoost [17]. Lilhore et al. combined CNN, Bi-LSTM, and EfficientNet-B0 to achieve high-accuracy classification [25].

Table 5 details frequently used ML models, their applications, strengths, limitations, and citations, providing a comprehensive overview of their roles in breast cancer detection.

### 2.2 Hybrid Model for Prediction

The hybrid models have been introduced as a radical solution to the issue of standalone ML models in breast cancer detection, as they can do what standalone models could not. The models are generally combinations of deep learning feature extractors, like CNNs, and more traditional classifiers or modern neural computing, being more accurate, robust, and understandable. Qasrawi et al. elaborated the hybrid model using DenseNet121 to extract features and an ensemble deep Random Vector-Functional Link Neural Network (edRVFL) that accomplished an excellent detection outcome in clinical practice [9]. The U-KAN model presented by Zhu et al. is a combination of U-Net based on CNN and KAN blocks, which is more accurate and interpretable in segmentation [11]. Essa et al. introduce a multi-stage hybrid model that converts biomarker values into image form to be classified by the ResNet50 model; thus, removing variability in the biomarker variable [15].

Twum et al. applied pre-trained CNNs such as ResNet50 and VGG16 as feature extraction and supervised classifiers, such as LightGBM and XGBoost, improving the results on ultrasound images [13]. Singh et al. offered a system that combines global descriptors of VGG16 with local ones that are being employed through a custom CNN, which are then categorized by a combination of SVM and Decision Trees, with a high accuracy [24]. Lilhore et al. created a hybrid model comprising CNN, Bi-LSTM, and EfficientNet-B0 to achieve accurate classification using the temporal dependence [25]. A new feature selection algorithm that included SHAP, RFE, and RF combined and optimized by PSO was proposed by Zhu et al. to be used for early detection [17]. Gaussian Process Hyperparameters in RF, GBT, and MLP models have been optimized using Genetic Algorithms to make them perform better on multi-modal data [16]. The BCDGAN framework presented by Veerlapalli and Dutta combined GANs with a hybrid classifier that consisted of InceptionResNetV2 and VGG16 and improved feature extraction [8]. Shen et al. transformed ResNet50 and VGG16 to whole-image classifiers with convolutional layers to test hybrid networks with diverse architecture [26].

Table 2 presents an overview of the hybrid models, their constituents, their uses, strengths, and limitations, and references.

**Table 2.** Hybrid Models in Breast Cancer Detection.

| Hybrid Model | Components | Application | Strengths | Limitations | Citation |
| --- | --- | --- | --- | --- | --- |
| DenseNet121 +<br>edRVFL | DenseNet121,<br>edRVFL | Classification | High accuracy,<br>robust | Device<br>variability | [9] |
| U-KAN | U-Net, KAN blocks | Segmentation | Improved<br>accuracy,<br>interpretability | Early-stage<br>adoption | [11] |
| ResNet50 + SVM | ResNet50, SVM | Classification | Robust,<br>interpretable | Biomarker<br>variability | [15] |
| VGG16 + Custom<br>CNN | VGG16, Custom CNN,<br>SVM, DT | Classification | Feature fusion,<br>high accuracy | Computational<br>complexity | [24] |
| CNN + Bi-LSTM +<br>EfficientNet-B0 | CNN, Bi-LSTM,<br>EfficientNet-B0 | Classification | Temporal<br>dependency<br>modeling | Real-time<br>challenges | [25] |
| RF + SHAP + RFE +<br>PSO | RF, SHAP, RFE, PSO | Classification | Enhanced feature<br>selection | High<br>computational<br>cost | [17] |
| BCDGAN | GAN,<br>InceptionResNetV2,<br>VGG16 | Classification | Enhanced feature<br>extraction | High<br>computational<br>cost | [8] |
| GA-optimized RF,<br>GBT, MLP | RF, GBT, MLP, GA | Classification | Optimized<br>performance | Single database | [16] |
| ResNet50 +<br>LightGBM | ResNet50, LightGBM | Classification | Transfer learning,<br>efficient | Risk of<br>overfitting | [13] |
| VGG16 +<br>Convolutional Layers | VGG16, Additional<br>Conv Layers | Classification | Whole-image<br>classification | Image<br>downsizing<br>issues | [26] |
| ResNet50 + Fuzzy NN | ResNet50, Fuzzy NN | Classification | Novel feature<br>engineering | Biomarker<br>variability | [15] |
| MobileNetV2 +<br>XGBoost | MobileNetV2,<br>XGBoost | Classification | Lightweight,<br>robust | Dataset<br>variability | [13] |
| InceptionResNetV2 +<br>VGG16 | InceptionResNetV2,<br>VGG16 | Classification | Enhanced feature<br>extraction | Single dataset | [8] |
| RF + CNN | RF, CNN | Multi-modal<br>Classification | Integrates diverse<br>data | Small sample<br>size | [22] |
| SVM + Custom CNN | SVM, Custom CNN | Classification | Robust<br>classification | Computational<br>complexity | [24] |
| End2End + Grad-CAM | End2End, Grad-CAM | Classification | Saliency insights | Interpretability challenges | [21] |
| DMV-CNN + Grad-CAM | DMV-CNN, Grad-CAM | Classification | Multi-view analysis | Small sample size | [21] |
| GMIC + Grad-CAM | GMIC, Grad-CAM | Classification | Global awareness | Interpretability issues | [21] |
| GLAM + Grad-CAM | GLAM, Grad-CAM | Classification | Local-global integration | Limited sample size | [21] |
| RF + MLP | RF, MLP | Classification | Multi-modal integration | Single database | [16] |

### 2.3 Kolmogorov-Arnold Networks (KAN) Applications

Kolmogorov-Arnold Networks (KANs) are a revolutionary ML paradigm, motivated by the Kolmogorov-Arnold representation theorem, that asserts that any multivariate continuous function can be decomposed as a composition of univariate functions and summation. In contrast to conventional deep neural networks, whose activation functions (e.g., ReLU) and weight matrices are fixed, KANs use trainable univariate functions and can describe relationships in complex and non-linear ways, which can be interpreted, and this makes KANs more general and flexible than conventional neural networks. Such flexibility is of great importance in the field of medical imaging, where there can be a lot of variability in the dataset, owing to disparities in imaging apparatus, patient demographics, and presentation of disease. The simpler model complexity of KANs that applies to functional decomposition may lead to decreased computational requirements and performance preservation or enhancement, which is better suited in clinical situations.

Zhu et al. suggested a U-KAN model in which KAN blocks were combined with a U-Net structural framework in the segmentation of breast cancer tumor lesions, containing the advantage of good accuracy and interpretability [11]. U-KAN generates tokenized KAN blocks in an encoder-decoder architecture, showing segmentation efficiencies better than the conventional U-Net and U-Net++ designs in identifying intricate margins of the lesion in mammography. This is because of the capability of a KAN to learn feature representations adaptively, which is essential in dealing with the disparity in medical pictures. Although KANs are still at an embryonic stage of use, they show promise to ease the model architecture and enhance its explainability it which would make medical imaging tasks such as segmentation more viable to KANs. Further studies may consider KANs as a new opportunity in classification or in combination with other deep learning models to improve performance even more, which is established by the experimental results of the works of, e.g., Zhu et al. [11].

Table 3 gives an account of the comparison of KAN-based models and other models in segmentation and classification in terms of their application, strengths, products, limitations, and references.

**Table 3.** KAN and Related Models for Breast Cancer Detection.

| Model | Architecture | Application | Strengths | Limitations | Citation |
| --- | --- | --- | --- | --- | --- |
| U-KAN | U-Net + KAN blocks | Segmentation | High accuracy, interpretability | Early-stage adoption | [11] |
| U-Net | Encoder-Decoder CNN | Segmentation | Established, widely used | Less flexible than KANs | [11,14] |
| U-Net++ | Enhanced U-Net | Segmentation | Improved feature aggregation | Higher complexity | [11] |
| MobileNetV2 U-Net | MobileNetV2 + U-Net | Segmentation | Lightweight, efficient | Limited by nuclei information | [19] |
| Mask-RCNN | CNN-based | Segmentation | Effective for auto-cropping | Manual cropping needed | [20] |
| YOLOv5 | CNN-based | Detection | Fast, accurate detection | Device variability | [9] |
| MedSAM | Segmentation Model | Segmentation | Specialized for medical images | Limited dataset diversity | [9] |
| Faster R-CNN | CNN-based | Detection | Robust object detection | Computationally intensive | [14] |
| ResNet50 | Deep CNN | Classification | Deep feature extraction | High computational cost | [6,13,15] |
| VGG16 | Deep CNN | Classification | Simple, transfer learning | Large memory footprint | [8,20,26] |
| InceptionV3 | Deep CNN | Classification | Multi-scale features | Complex architecture | [7] |
| DenseNet121 | Deep CNN | Classification | Efficient feature reuse | Large dataset needs | [9] |
| EfficientNet-B0 | Deep CNN | Classification | Balanced efficiency | Real-time applicability | [25] |
| InceptionResNetV2 | Deep CNN | Classification | Enhanced feature extraction | High computational cost | [8] |
| Bi-LSTM | Recurrent Neural Network | Classification | Temporal dependency modeling | Real-time challenges | [25] |
| Random Forest | Ensemble Classifier | Classification | Interpretable, efficient | Limited to images | [10,22,25] |
| SVM | Linear Classifier | Classification | Robust for tabular data | Feature scaling sensitivity | [10,12,23] |
| Logistic Regression | Linear Classifier | Classification | Simple, interpretable | Limited for complex data | [10,12,23] |
| XGBoost | Gradient Boosting | Classification | High performance | Requires tuning | [10,13] |
| LightGBM | Gradient Boosting | Classification | Efficient, scalable | Dataset variability | [13,17] |

### 2.4 Gaps in Literature

In spite of all the progress, the research on ML in breast cancer detection areas suggests that there still exist several research gaps that hamper clinical implementation. One of these weaknesses is the attraction to small or single-institutional data, which affects the model’s generalizability, strength, and stability. As it was mentioned by Fornvik et al., the small cohort and the single equipment vendor became the limitations of their study, and they had to validate their 3D ResNet model using larger and multi-center datasets [6]. By the same token, Solorzano et al. emphasized a very low n of 587 WSIs [7], and Sinjanka et al. remarked on limitations due to a narrow, local population [22]. Model robustness is further posed by variability in image devices, such that Qasrawi et al. and Twum et al. noted discrepancies in using various equipment [9,13].

The fact that deep learning models are commonly referred to as black boxes is still a major barrier to interpretability. Fornvik et al. and Pertuz et al. applied partial solutions to Grad-CAM in studying decision-making, and these approaches are not sufficient to explain all aspects of a complex process [6,21]. Balkenende et al. highlighted the lack of studies on nuclear medicine, the necessity to have a wide range of data on ultrasound and MRI, and the solution of legal and ethical concerns on clinical implementation [14]. There is a lack of exploration regarding the multi-modal data integration, e.g., clinical, genomic, or lifestyle factors, to obtain comprehensive insights to use as diagnostics. As Essa et al. indicated, the available biomarker data were variable and likely to reduce the generality of their model [15], and Rabiei et al. pointed out the absence of genetic data [16].

The other issue is the computational complexity, especially with deep learning algorithms such as ResNet50 and VGG16 that demand many resources, making them hard to implement in resource-restricted environments [12,13]. According to Shen et al. and Veerlapalli and Dutta, there were problems with downsizing of images and significant computational costs of GAN-based models, respectively [8,26]. Huang et al. outlined barriers to making out distinctions of well-differentiated tumor cells because they utilize information at the nuclei level [19]. As Alotaibi et al. implied, it has been proposed that manual cropping can enhance the quality of the data [20]. Naji et al. and Ravi Kumar et al. noted the necessity to validate information on various databases to increase the level of generalizability [10,23]. It was emphasized by Singh et al. and Lilhore et al. that alternative feature selection and real-time implementation measures need to be examined [24,25]. According to Zhu et al., there were found to be difficulties in optimizing the processing parameters of varying datasets [17].

Table 4 outlines key research gaps, their impact, proposed solutions, and citations.

**Table 4.** Research Gaps in Breast Cancer Detection Literature.

| **Research Gap** | **Impact** | **Proposed Solutions** | **Citation** |
| --- | --- | --- | --- |
| Small/Single-Institution<br>Datasets | Limited generalizability | Multi-center studies, larger<br>datasets | [6,7,22,28–30,33,36] |
| Imaging Device Variability | Poor performance across<br>devices | Standardized protocols, diverse<br>datasets | [9,13] |
| Interpretability Challenges | Hinders clinical trust | Advanced explainability tools<br>(e.g., SHAP, LIME) | [6,21,29,34,35] |
| Limited Multi-Modal<br>Integration | Incomplete diagnostic<br>insights | Incorporate clinical, genomic<br>data | [15,22,29] |
| Computational Complexity | Resource constraints | Lightweight models,<br>optimization techniques | [12,13,31] |
| Limited Nuclei Information | Misleads AI in complex<br>tumors | Incorporate additional features | [19] |
| Manual Preprocessing Needs | Time-consuming, error-<br>prone | Automated preprocessing<br>pipelines | [20] |
| Single Database Reliance | Limited generalizability | Multi-database validation | [10,23] |
| Parameter Optimization<br>Challenges | Suboptimal performance | Adaptive parameter selection | [11] |
| Real-Time Implementation<br>Issues | Hinders clinical adoption | Optimize for low-power devices | [25,31] |
| Image Downsizing | Loss of detail | High-resolution processing | [26] |
| Lack of Genetic Data | Limited diagnostic scope | Integrate genomic data | [16] |
| Sparse Nuclear Medicine<br>Research | Underdeveloped modality | Expand research focus | [14] |
| Legal/Ethical Issues | Delays clinical<br>implementation | Develop regulatory frameworks | [14,27,34] |
| Limited Feature Selection<br>Methods | Suboptimal feature sets | Explore internal methods,<br>wrappers | [24] |
| Overfitting on Small Datasets | Poor generalization | Data augmentation,<br>regularization | [13] |
| Biomarker Variability | Affects model generality | Robust feature engineering | [15] |
| High Computational Cost of<br>GANs | Resource-intensive | Optimize GAN architectures | [8] |
| Small Regional Samples | Limited representativeness | Diverse, global datasets | [22] |
| Lack of Pathological<br>Confirmation | Relies on BI-RADS | Incorporate pathology data | [26] |
| Lack of Temporal<br>Comparison | Misses key radiologist<br>workflow step | Develop models capable of<br>longitudinal analysis | [27] |
| Low Sensitivity in Dense<br>Breasts | Missed diagnoses | Multi-modal approaches (e.g.,<br>DBT, CESM) | [31] |
| Subjective Breast Density<br>Assessment | High inter-observer<br>variability | Objective, quantitative ML<br>methods | [33] |
| Suboptimal Feature Fusion | Does not leverage full model<br>strengths | Develop more advanced fusion<br>techniques | [32] |
| Publication Bias | Skewed view of model<br>performance | Encourage publication of all<br>results, meta-analyses | [34] |
| Unclear Clinical Role | Ineffective model<br>optimization | Clearly define intended use<br>(e.g., screening, diagnosis) | [34] |
| Annotation<br>Quality/Consistency | Poor model performance | Standardized annotation<br>protocols | [35] |
| Data Privacy and Security | Hinders data sharing and<br>collaboration | Develop secure data-sharing<br>frameworks | [35] |
| Scarcity of High-Quality<br>Data | Limits robust model training | Create large, diverse, well-<br>annotated public/private datasets | [34,35] |

## 3. Methodology

The chapter presents the research method to be used in testing eight machine learning (ML) models to help in the detection of breast cancer based on mammogram imaging. This research capitalizes on the Mammogram Mastery dataset to identify the performance, computational economy, and clinical translation of Convolutional Neural Network (CNN), Kolmogorov-Arnold Network (KAN), k-Nearest Neighbors (kNN), Support Vector Machine (SVM) and XGBoost, Random Forest, Naive Bayes, and a new-fangled Hybrid model. The methodology is arranged in three subsections: firstly Dataset, secondly Machine Learning Models, and thirdly Evaluation Metrics.

### 3.1 Dataset

This work employs the Mammogram Mastery dataset published in April 2024 by Iraq-Sulaymaniyah with 745 original mammogram images and 9,685 augmented images, as curated and labeled images by specialist clinicians, thus with very high reliability and accuracy [18]. This is a dataset that has been sourced in an area not currently represented within a global medical dataset, hence the dataset gives a rather rare insight into the nature of breast cancer and aids in the fabrication of diagnostic tools that can be used across a wide variety of people. The increased images are created using such techniques as random rotations (up to 20 degrees), horizontal flips, and zooming (1020 percent range), and make the data more versatile and allowing models to learn under diverse imaging conditions to minimize the risks of overfitting [18].

Images undergo preprocessing to uniformize and make compatible with ML models, resized to 128x128 pixels to standardize the size of the input, normalized to a range of intensity [0,1] to decrease variation in the intensity of pixels, and augmented to increase the amount and variety of them. The dataset was divided into 80% training and 20% testing datasets with a similar proportion of cancerous and non-cancerous cases in each. Such a split allows positive modeling training, optimizing hyperparameters, and unbiased estimation of performance standards that adhere to the best practice in medical imaging studies [13,14].

### 3.2 Machine Learning Models

Eight ML models were tested, and each was chosen because of their unique binary classification of mammograms. A brief description of each of the models and their configuration, architecture, and training follows below:

#### 3.2.1 Convolutional Neural Network (CNN) Model

The Convolutional Neural Network (CNN) in the notebook presented corresponds to a deep learning architecture that is particularly used in analyzing medical images, as it performs the classification of medical images, such as breast cancer detection. Building this model is performed with torch.nn.Module and commonly includes various forms of layers. The design of the convolution layers (nn.Conv2d) is ubiquitous as they involve the application of the learnt filters in extracting the hierarchical features of the images given as input, commonly succeeded by Batch Normalization (nn.BatchNorm2d) in stabilizing the training and ReLU activation functions (nn.ReLU) in the induction of non-linearity. To make the model invariant to small translations and decrease the complexity of computation, the spatial dimensions in the feature maps are then pooled using pooling layers, e.g., Max Pooling (nn.MaxPool2d). Following a succession of such blocks, the acquired 2D feature maps are converted to 1D vectors and run through one or more fully connected (linear) layers (nn.Linear), accomplishing the last stage of classification based on obtained high-level features. It may also involveDropout layers to guard against overfitting.

It consists of 3 layers, namely, convolutional, pooling, and fully connected layers, and represents 8,482,369 parameters. There are 32 filters in the first layer (3x3 kernels), the second layer is max-pooling and dropout (0.5 rate) to avoid overfitting, and in the last layer, there is a dense layer with sigmoid activation since it requires a binary output. The CNN architecture was trained on ImageNet and fine-tuned on the Mammogram Mastery data set, and the learning rate as well as the batch size were optimized through the use of grid search [6,24]. Fig 2 shows the block diagram of the CNN model architecture.

**Fig 2.**
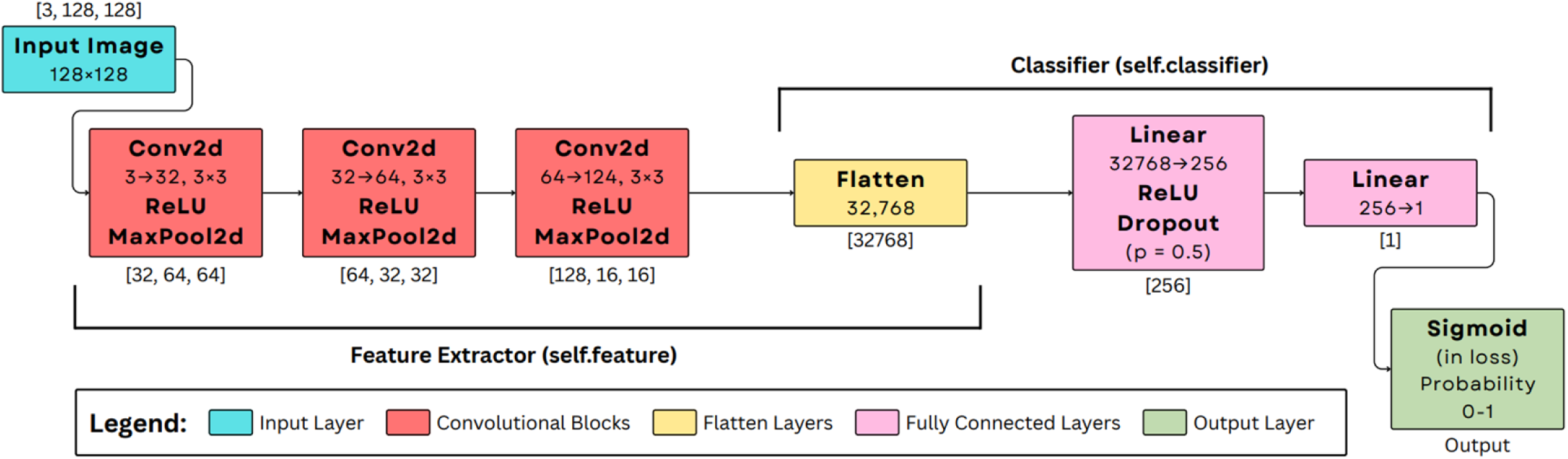
Used CNN Model Architecture.

#### 3.2.2 Kolmogorov-Arnold Networks (KAN) Model

KAN model is an alternative to the conventional architecture of a neural network because it introduces the activation functions at the edges (connections), instead of the nodes. This network design that learns explicit mathematical functions on connections is implemented with the help of the pykan library and will make the implemented model more tractable. The width of the architecture or layers defines the number of neurons in each layer as per a multi-layer perceptron. Nevertheless, the crux of the matter is that the links between the neurons in KAN have each such a spline that is to be learnt. This design distinguishes upon the fact that running KANs might find and express encoded symbolic relationships to the data, and this could be really useful in grokking the decision-ratio in tricky apps such as breast cancer detection, giving a level of transparency that black-box models lack. The architecture of the proposed KAN model is illustrated in Fig 3.

**Fig 3.**
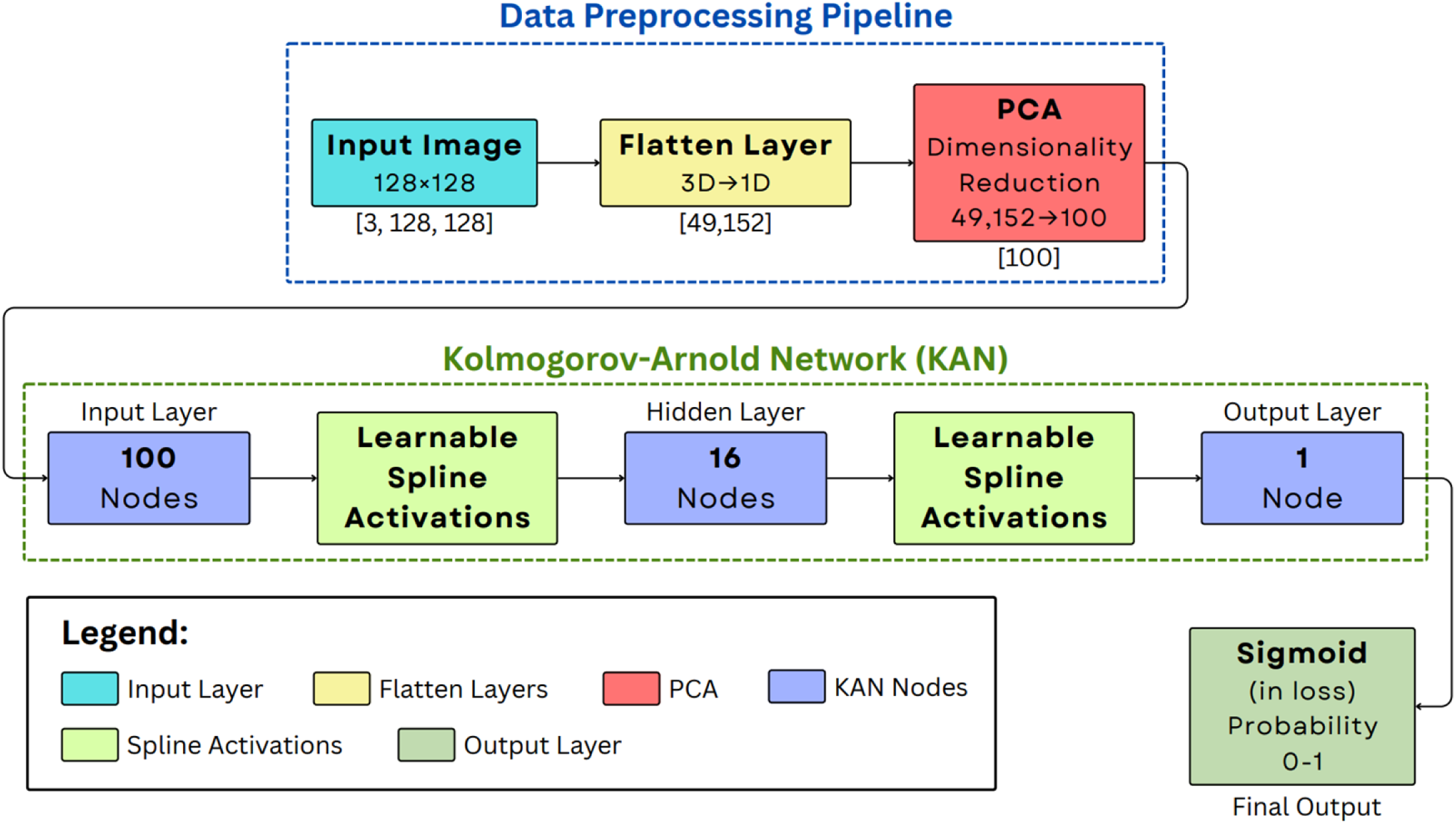
Used KAN Model Architecture.

It is a new architecture containing 22,624 parameters and uses spline-based function approximation to model the complex relationships, and requires fewer parameters than ordinary neural networks [11]. KAN was set up in the approach that consisted of a single hidden layer, but the functions were of B-spline, and the model parameters were selected to balance performance and understandability. The hyperparameters, such as spline order, grid size, were optimized to find a balance between accuracy and computational cost.

#### 3.2.3 k-Nearest Neighbors (kNN) Model

K-Nearest Neighbors (kNN) model, instantiated by KNeighborsClassifier in sklearn.neighbors is a non-parametric and instance-based learning algorithm. As opposed to other models, kNN does not involve finding an explicit function during a learning process; to a large extent, it postpones calculations until it can make a prediction. Breast cancer detection: To detect breast cancer, say we have a new picture (or its extracted features), then the kNN method finds the k nearest data points (i.e., the neighbors) in the training set according to some selected distance measure, e.g., Euclidean distance. The category of the new image can simply be determined as the most common of these k nearest neighbors, thus labeling it with this category label. The kNN algorithm is extremely sensitive to the parameters of k (size of the neighborhood) and the distance measure, as these parameters directly affect the definition of the neighborhood and, in turn, determine the result of the classification.

Such a model has been set with 5 neighbors and makes use of 29,294,592 training points (obtained by flattening image features). The kNN model uses the Euclidean distance to classify, which in the case of small data is applicable, and the image data is computationally expensive because it uses distance calculations in the process [12,23]. The performance was maximized through the selection of the number of neighbors using cross-validation.

#### 3.2.4 Naive Bayes Model

Naive Bayes, in particular, GaussianNB, which is a sklearn.naive_bayes method seems to act on the assumption of Bayes because it assumes the strong premise that all characteristics of a given class are going to be independent. In a breast cancer detection application, this model would mean that the probability distribution of each feature (e.g., image pixel values or extracted features) will be learned in both classes; i.e., a Gaussian distribution will be learned in continuous data. In prediction, those distributions are used to compute the probability that a new datum belongs to each class, and the maximum probability class is chosen. It is rather simple and fast, but its activity may be restricted when a lack of independence of assumption takes place in the real data.

It is a Gaussian Naive Bayes model with 196,608 features (49,152 pixels, 2 classes), and there are also assumptions that the features are independent. This model has a low computation cost but can be problematic when dealing with the intricate spatial relationships of mammogram images [23]. Variance smoothing was set up to manage numeric stability in the course of training.

#### 3.2.5 Random Forest Model

Random Forest model is an ensemble learning method that is based on constructing a large number of decision trees to be used during the training. In the case of breast cancer detection, it implies that rather than using one decision tree with limited accuracy and overfitting issues, the model will utilize multiple trees to predict the issue and increase accuracy and reduce overfitting. A random subset of the training data is used to construct each individual tree in the forest. This is known as bootstrapping. At each internal node of the trees, only a random subset of the features is taken into consideration. Such randomness assists in decorrelating the trees and therefore the ensemble becomes robust. Every tree in the forest casts a vote, and the decision is taken by simple majority among the trees, so that the wisdom of the crowds is applied in generating a more reliable decision.

It is a group of 100 decision trees whose average depth is 8.89 (approximately 4,542 nodes comprise the group). Bagging as well as randomness of the features is used in the model in order to minimize overfitting; hence, the model is robust in handling the pre-extracted features [10,12]. Tree depth and number of trees were the hyperparameters that were optimized using grid search.

#### 3.2.6 Support Vector Machine (SVM) Model

Support Vector Machine (SVM) model, to be specific, the model of SVC or Support Vector Classifier on sklearn.svm, is a strong discriminative classifier that attempts to seek an optimal hyperplane in its N-dimensional space (which is the number of features) to categorically differentiate the data points into different classes. With regard to identifying breast cancer, the SVM attempts to establish a precise distinction between cancer and non-cancer cases. In instances where data cannot be linearly separated, the method used within SVM takes into consideration the so-called "kernel trick". This method tacitly dimensionally projects the input features into a higher-dimensional feature space in which they could be partitioned linearly. Linear kernel and Radial Basis Function (RBF) kernel, which are used in simple linear and complex, non-linear relationships, respectively, are common kernel functions. SVM aims not only to classify the classes by using a separating hyperplane, but also to use as big a margin as possible between the same hyperplane and the nearest data points (called support vectors), so that it helps to improve the capacity of generalization of the model.

Meaning that this model will have a linear kernel and 257 support vectors, hence this model becomes an SVM model that will work on flattened image features, leading to 12,632,322 parameters. A linear kernel was selected due to its ease of use and performance in high-dimensional spaces that fit pre-extracted features of mammograms [12,23]. The regularization parameter C and other hyperparameters were optimized by grid search over the validation set to maximize cross-validation.

#### 3.2.7 XGBoost Model

XGBoost model is a highly powerful and efficient ensemble learning method, belonging to the gradient boosting approach, that is executed as XGBClassifier under the xgboost library. In breast cancer detection by XGBoost, a number of weak models that are usually prediction trees are constructed sequentially, with a later tree designed to fix the mistakes made by an earlier tree, and then the next tree is created to fix the mistakes of the prior tree. It stands out due to some optimizations: it supports regularization terms (L1 and L2), avoiding overfitting and thus is more resistant; it is suitable for missing values directly (usually by setting them to some target value); and uses advanced pruning and parallel training to speed up training and gain better results. This set of properties enables XGBoost to be very accurate and efficient, and therefore, it tends to be used in solving structured data prediction problems.

It is a gradient boosted tree model, and the hyperparameters are default and optimized for tabular features of mammograms. XGBoost is based on iterative boosting to reduce the prediction errors, which provides good results on structured data [10,13]. Such parameters as the learning rate or the maximum depth were optimized to avoid overfitting and achieve better accuracy.

#### 3.2.8 Hybrid Model

The Hybrid Model is a sophisticated system that integrates the best features of various machine learning paradigms to produce better results due to the possibility of better breast cancer diagnosis. This specific hybrid architecture combines a pre-trained Convolutional Neural Network (to be more precise, ResNet18), Gaussian Naive Bayes, and a Random Forest. ResNet18 is a robust feature extractor and has a deep architecture as well as pre-trained weights, which generate feature-rich and high-level features on the basis of the reliance on input medical images. Such deduced features are then piped into a Gaussian Naive Bayes classifier and a Random Forest classifier session, permitting these different models to learn from the same authoritative characterization. The predicted output of the last combination of the hybrid system is obtained by ensembling the output of all three models (pre-issued CNN, Naive Bayes, and Random Forest). Therefore, this ensemble may use a weighted average or voting scheme of the majority, thus applying the various decision-making processes of each component model to increase accuracy and resistance to false positive or negative inference.

It is a new architecture, which integrates CNN feature extraction (pre-trained on ImageNet) and an ensemble classifier (Random Forest and XGBoost), and the number of their parameters is 35,109,377. CNN is used to extract deep features on mammogram images, and the resulting features are used in an ensemble of 50 Random Forest trees and an XGBoost classifier, and the predictions are combined through soft voting. This type of hybrid model is essentially combining the power of deep learning in feature extraction and the power of the ensemble notion in resilient classification [9,24]. Grid search on the validation set was done to optimize hyperparameters (such as CNN learning rate and ensemble weights). The proposed Hybrid model architecture is illustrated in Fig 4.

**Fig 4.**
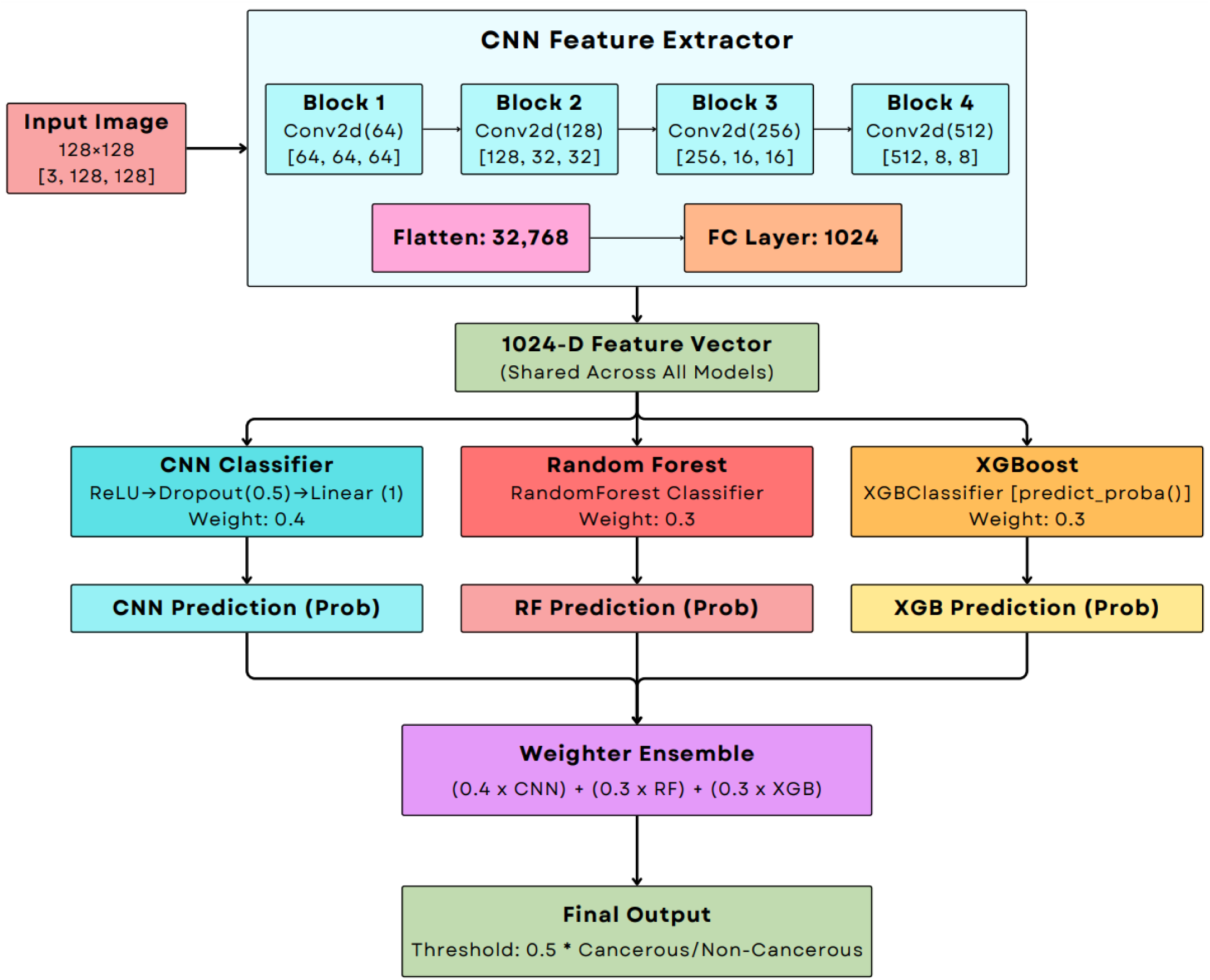
Proposed Hybrid Model Architecture.

To provide scalability, all models were trained on a GPU cluster (NVIDIA RTX 3090, 24GB VRAM), and early stopping and learning rate scheduling were used to avoid overfitting. Real-time augmentation of training data was used to increase robustness, and hyperparameters were optimized with 5-fold cross-validation on the validation set to achieve an ideal performance.

### 3.3 Evaluation Metrics

In an attempt to develop a thorough evaluation of the models and their future clinical implementation, the following metrics were utilized, according to the outline:

**Accuracy**: The number of accurate predictions (true positives and true negatives) is estimated based on the number of overall predictions that give a degree of model accuracy.

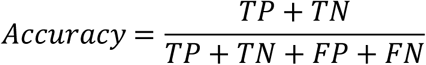

where:

- **TP (True Positives):** The count of non-cancerous cases predicted accurately.
- **TN (True Negatives):** The count of cancerous cases predicted accurately.
- **FP (False Positives):** The count of cancerous cases that are falsely predicted to be non-cancerous.
- **FN (False Negatives):** The count of non-cancerous cases that are falsely predicted to be cancerous.

The concept of accuracy is mostly applicable to balanced data, such as Mammogram Mastery, but on the other hand, it might not be very revealing in the case of an imbalanced dataset [14].

**F1 Score**: The harmonic mean of precision and recall, calculated as:

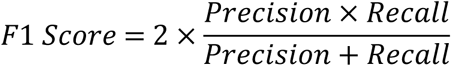

where:

- **Precision** measures the accuracy of the positive predictions.

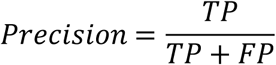
- **Recall** (also known as Sensitivity or True Positive Rate) measures the ability of the model to identify all relevant instances.

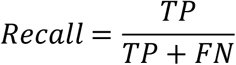

This metric is critical for imbalanced datasets, as it balances the trade-off between correctly identifying cancerous cases (recall) and minimizing false positives (precision), ensuring robust performance in medical diagnostics [9].

**ROC AUC Score**: The area under the receiver operating characteristic curve, measuring the model’s ability to distinguish between cancerous and non-cancerous classes across various classification thresholds. It is the area under the curve that plots the True Positive Rate (Recall) against the False Positive Rate at various classification thresholds.

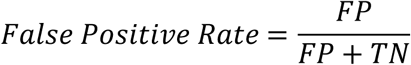

A higher ROC AUC indicates better discriminative performance, crucial for reliable diagnostics [13]. An AUC score of 1.0 represents a perfect classifier, while a score of 0.5 suggests no discriminative ability, equivalent to random guessing.

**Log Loss**: Log Loss, also known as logistic loss or cross-entropy loss, quantifies the accuracy of a classifier by penalizing false classifications. It measures the uncertainty of the predictions based on how much they vary from the actual labels. It is calculated as:

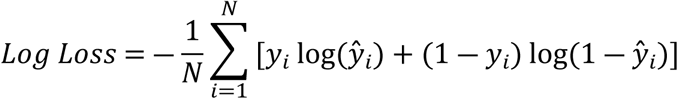

where:

- *N* is the total number of samples.
- *y*_*i*_ is the true label of the *i*-th sample (0 for non-cancerous, 1 for cancerous).
- *ŷ*_*i*_ is the predicted probability of the *i*-th sample being cancerous.

Lower log loss indicates more confident and accurate predictions, essential for clinical trust [11].

- **Inference Time**: The average time (in seconds) to process a single mammogram image, critical for real-time clinical applications where rapid diagnostics are necessary to integrate into busy workflows [13,25].
- **Training Time and Model Complexity**: Training time (in seconds) measures the computational cost of model training, while model complexity (e.g., number of parameters, support vectors, or nodes) assesses scalability and feasibility for deployment in resource-constrained environments, such as community clinics [9,13].

These metrics were computed on the test set to ensure unbiased evaluation, with results aggregated to compare model performance and trade-offs. The use of multiple metrics ensures a holistic assessment, addressing both predictive accuracy and practical considerations for clinical deployment.

## Chapter 4: Results

### 4.1 Performance Comparison

The performance of the Support Vector Machine (SVM), Convolutional Neural Network (CNN), a Hybrid model, Kolmogorov-Arnold Network (KAN), K-Nearest Neighbors (KNN), Naive Bayes, Random Forest, and XGBoost models was evaluated. A summary of the key performance metrics is presented in Table 6.

**Table 5.**
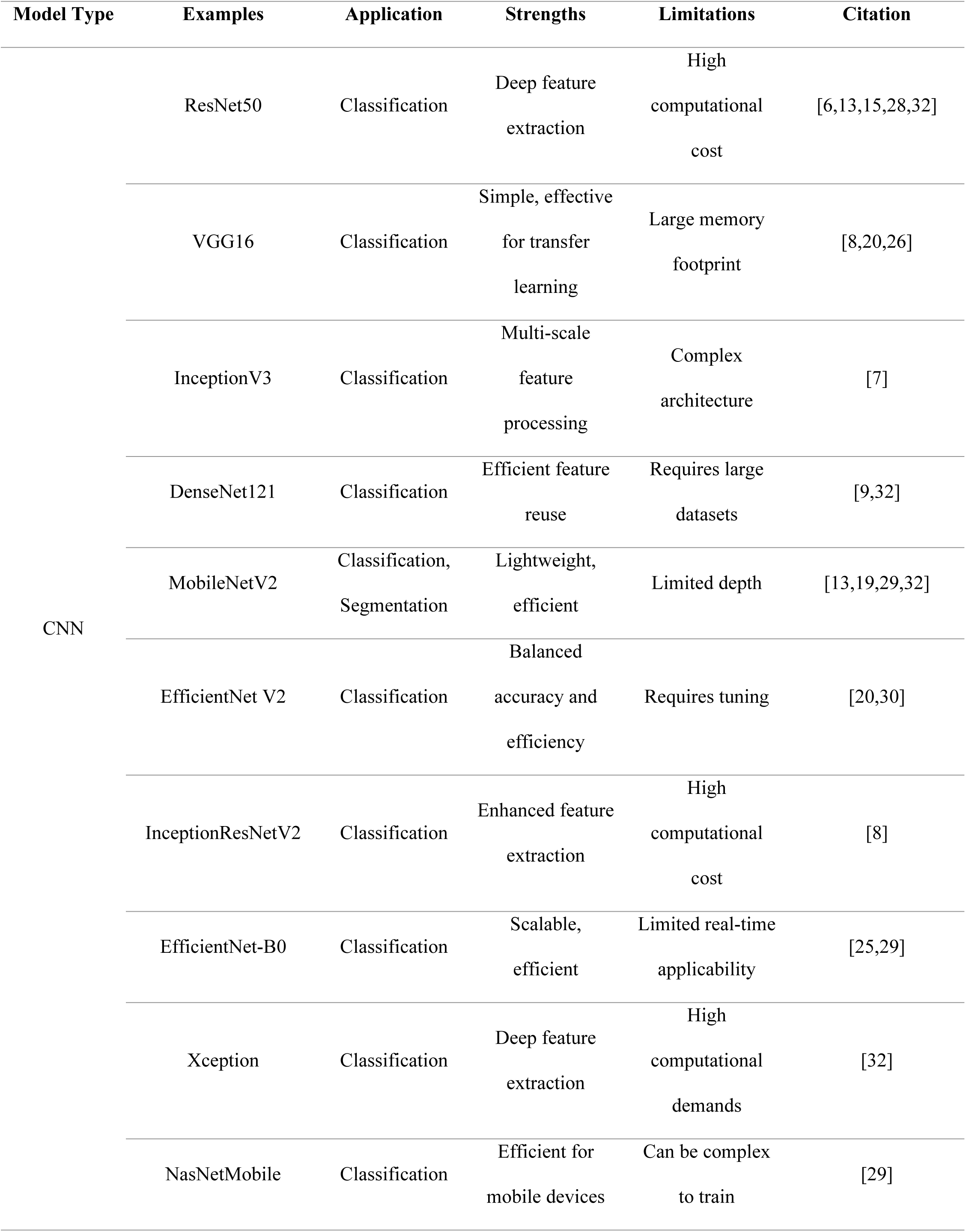
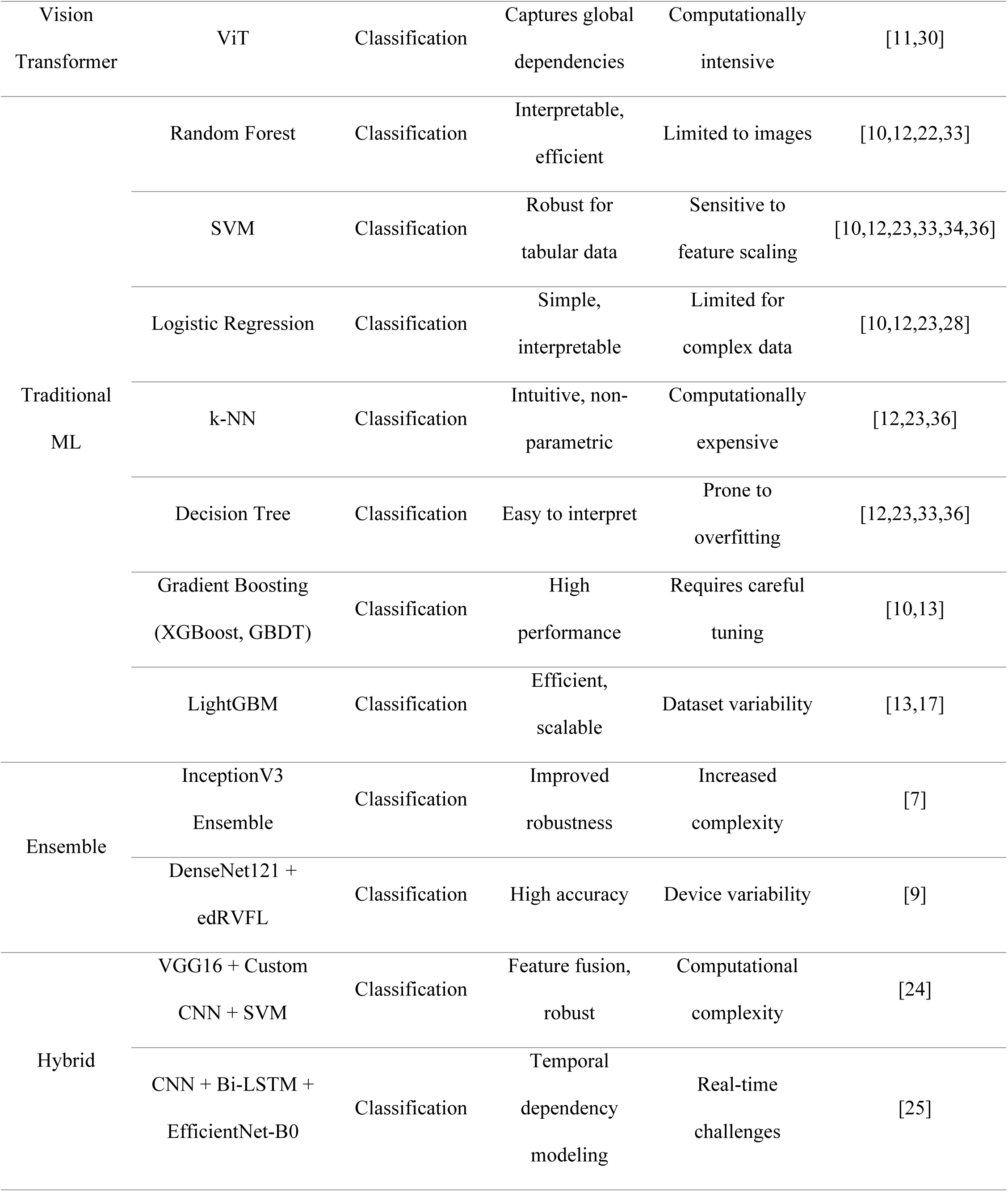
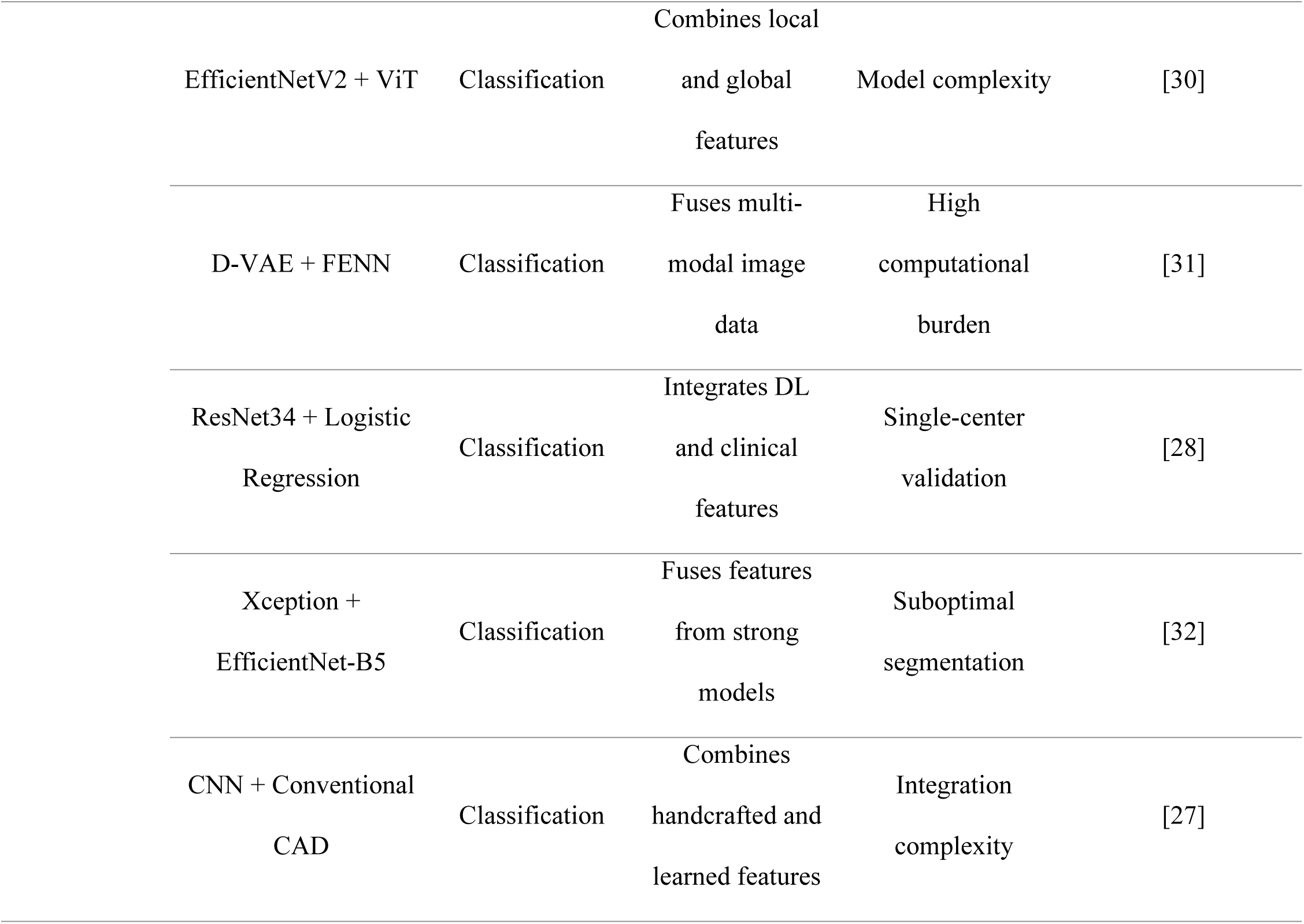
Commonly Used Models in Breast Cancer Detection.

| Model Type | Examples | Application | Strengths | Limitations | Citation |
| --- | --- | --- | --- | --- | --- |
| CNN | ResNet50 | Classification | Deep feature extraction | High computational cost | [6,13,15,28,32] |
|  | VGG16 | Classification | Simple, effective for transfer learning | Large memory footprint | [8,20,26] |
|  | InceptionV3 | Classification | Multi-scale feature processing | Complex architecture | [7] |
|  | DenseNet121 | Classification | Efficient feature reuse | Requires large datasets | [9,32] |
|  | MobileNetV2 | Classification, Segmentation | Lightweight, efficient | Limited depth | [13,19,29,32] |
|  | EfficientNet V2 | Classification | Balanced accuracy and efficiency | Requires tuning | [20,30] |
|  | InceptionResNetV2 | Classification | Enhanced feature extraction | High computational cost | [8] |
|  | EfficientNet-B0 | Classification | Scalable, efficient | Limited real-time applicability | [25,29] |
|  | Xception | Classification | Deep feature extraction | High computational demands | [32] |
|  | NasNetMobile | Classification | Efficient for mobile devices | Can be complex to train | [29] |
| Vision Transformer | ViT | Classification | Captures global dependencies | Computationally intensive | [11,30] |
| Traditional ML | Random Forest | Classification | Interpretable, efficient | Limited to images | [10,12,22,33] |
|  | SVM | Classification | Robust for tabular data | Sensitive to feature scaling | [10,12,23,33,34,36] |
|  | Logistic Regression | Classification | Simple, interpretable | Limited for complex data | [10,12,23,28] |
|  | k-NN | Classification | Intuitive, non-parametric | Computationally expensive | [12,23,36] |
|  | Decision Tree | Classification | Easy to interpret | Prone to overfitting | [12,23,33,36] |
|  | Gradient Boosting (XGBoost, GBDT) | Classification | High performance | Requires careful tuning | [10,13] |
|  | LightGBM | Classification | Efficient, scalable | Dataset variability | [13,17] |
| Ensemble | InceptionV3 Ensemble | Classification | Improved robustness | Increased complexity | [7] |
|  | DenseNet121 + edRVFL | Classification | High accuracy | Device variability | [9] |
| Hybrid | VGG16 + Custom CNN + SVM | Classification | Feature fusion, robust | Computational complexity | [24] |
|  | CNN + Bi-LSTM + EfficientNet-B0 | Classification | Temporal dependency modeling | Real-time challenges | [25] |

|  |  |  |  |  |
| --- | --- | --- | --- | --- |
| EfficientNetV2 + ViT | Classification | Combines local<br>and global<br>features | Model complexity | [30] |
| D-VAE + FENN | Classification | Fuses multi-modal image<br>data | High<br>computational<br>burden | [31] |
| ResNet34 + Logistic<br>Regression | Classification | Integrates DL<br>and clinical<br>features | Single-center<br>validation | [28] |
| Xception +<br>EfficientNet-B5 | Classification | Fuses features<br>from strong<br>models | Suboptimal<br>segmentation | [32] |
| CNN + Conventional<br>CAD | Classification | Combines<br>handcrafted and<br>learned features | Integration<br>complexity | [27] |

**Table 6.**
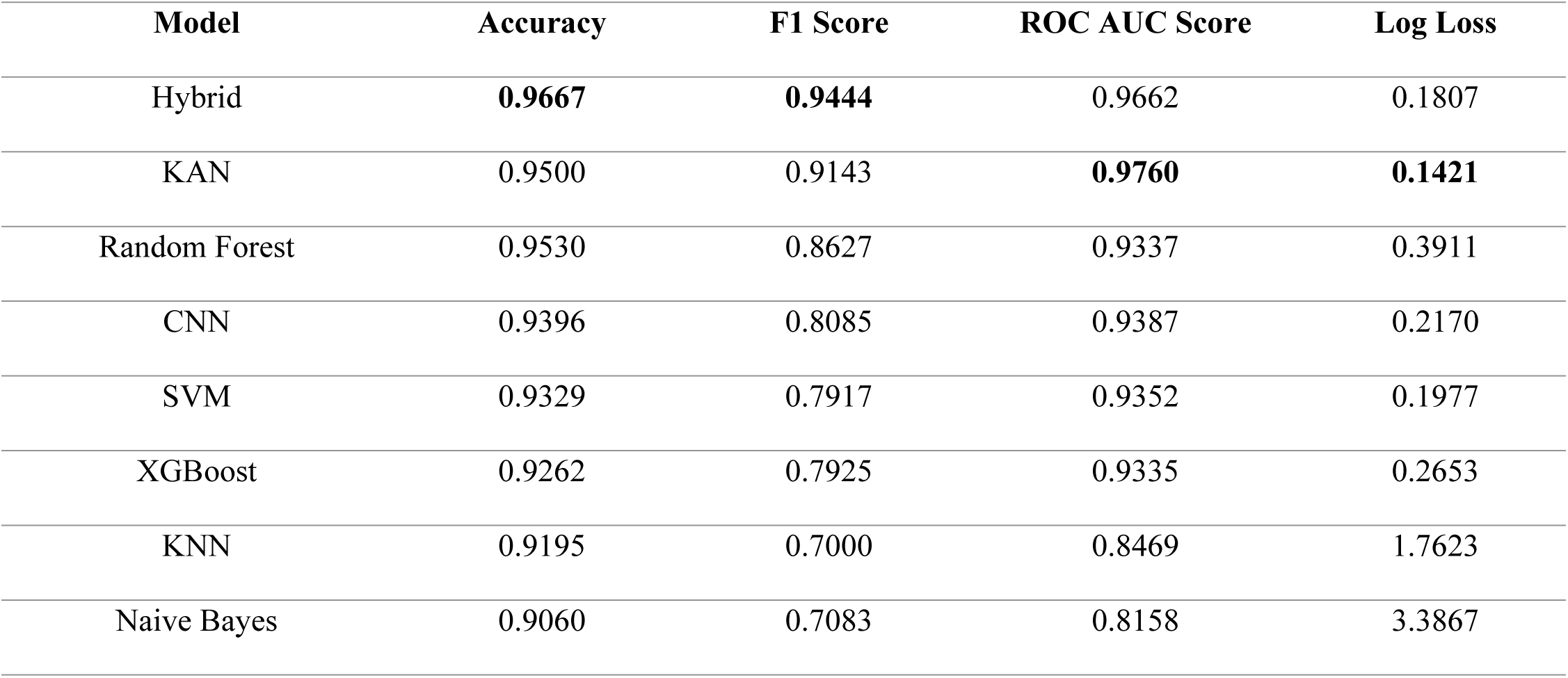
Performance Metrics of Evaluated Models.

| Model | Accuracy | F1 Score | ROC AUC Score | Log Loss |
| --- | --- | --- | --- | --- |
| Hybrid | 0.9667 | 0.9444 | 0.9662 | 0.1807 |
| KAN | 0.9500 | 0.9143 | 0.9760 | 0.1421 |
| Random Forest | 0.9530 | 0.8627 | 0.9337 | 0.3911 |
| CNN | 0.9396 | 0.8085 | 0.9387 | 0.2170 |
| SVM | 0.9329 | 0.7917 | 0.9352 | 0.1977 |
| XGBoost | 0.9262 | 0.7925 | 0.9335 | 0.2653 |
| KNN | 0.9195 | 0.7000 | 0.8469 | 1.7623 |
| Naive Bayes | 0.9060 | 0.7083 | 0.8158 | 3.3867 |

The results indicate a clear hierarchy concerning the performance of the models. The Hybrid model became the best option in regard to raw accuracy (0.9667) and F1 Score (0.9444). This reflects not merely a high rate of correct predictions, but also a better tradeoff between precision (what percent of those with positives were actually positive) and recall (what percent of actual positives were positively identified).

The newly proposed KAN model managed to display remarkable performance, with the greatest ROC AUC (0.9760) and the least Log Loss (0.1421). A good ROC AUC indicates an excellent performance in separating between the positive and negative classes at any classification level. Also, the value of the Log Loss score is quite low, which indicates that the probability estimates per hundred based on the KAN model are calibrated and are confident in their predictions, i.e., it is not just accurate, but is also sure of its accurate predictions.

Traditional models like KNN and Naive Bayes were way behind, however. Their poorer results in most of the measures imply that their prior assumptions and mechanisms could be too simplistic to depict the complex patterns occurring in complex medical images such as mammograms.

### 4.2 Computational Efficiency

In addition to the predictive accuracy, the degree to which a machine learning model can be effectively used in a clinical context is quite influenced by its computing requirements. Model development and retraining may be slowed down by long training times, and inference speeds may lag in high-throughput screening processes. Table 7 provides the profiles of the computational efficiency of the compared models.

**Table 7.** Computational Efficiency of Evaluated Models.

| Model | Training Time (seconds) | Average Inference Time (seconds) |
| --- | --- | --- |
| CNN | 1038.91 | **0.000012** |
| Hybrid | 1009.10 | 0.000047 |
| KAN | 328.99 | 0.322962 |
| XGBoost | 64.55 | 0.034322 |
| SVM | 49.56 | 0.019505 |
| Random Forest | 1.69 | 0.007408 |
| Naive Bayes | 0.23 | 0.000902 |
| KNN | **0.13** | 0.253339 |

The data shows an evident and anticipated trade-off between the model complexity, predictive performance, and computational cost. The best-performing deep learning models, CNN and the Hybrid model demanded initial investment of training time to the greatest degree, looking at approx. 1000 seconds each. But trained, these models are extraordinarily speedy at inference. KAN model offers a more peculiar profile, as its training time is moderate, and by far the slowest inference speed. On the other extreme, the Random Forest model stands out as an example of efficiency, given the fact that it takes a short time in training and is also faster in making inferences.

### 4.3 ROC Curve and Confusion Matrix Analysis

Although aggregate measures are valuable in describing an overall picture, they may conceal important information regarding the diagnostic behavior of a model. In this instance, a medical application, it would be necessary to know the particular errors made by the model.

#### 4.3.1 ROC Curve Analysis

The ROC curves show the trade-off with the True Positive Rate (TPR) against False Positive Rate (FPR) that provides a complete picture of how well a model can diagnose at any possible classification threshold. After comparing the Area Under the Curve (AUC) of all eight models, it can be said that the models fall into different categories of performance.

**Fig 5.**
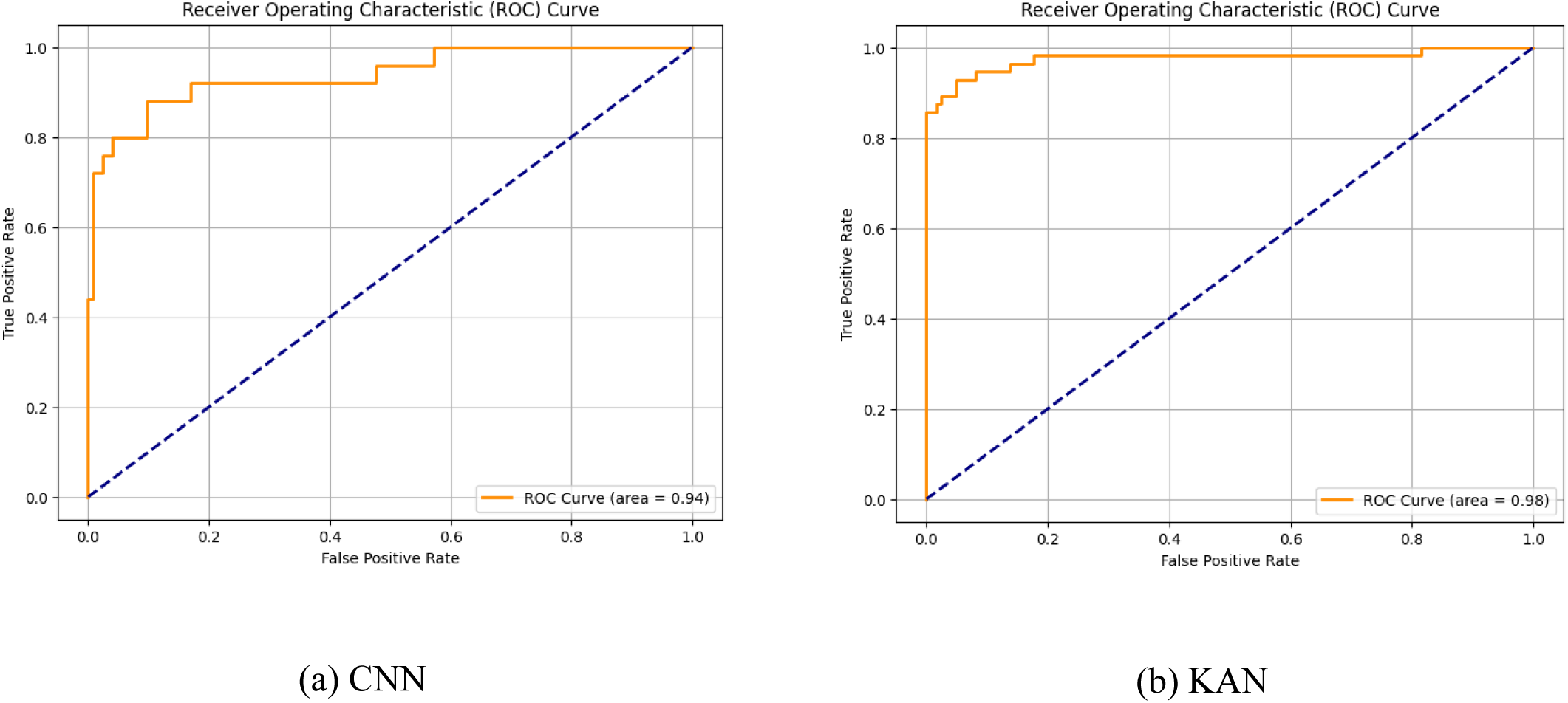
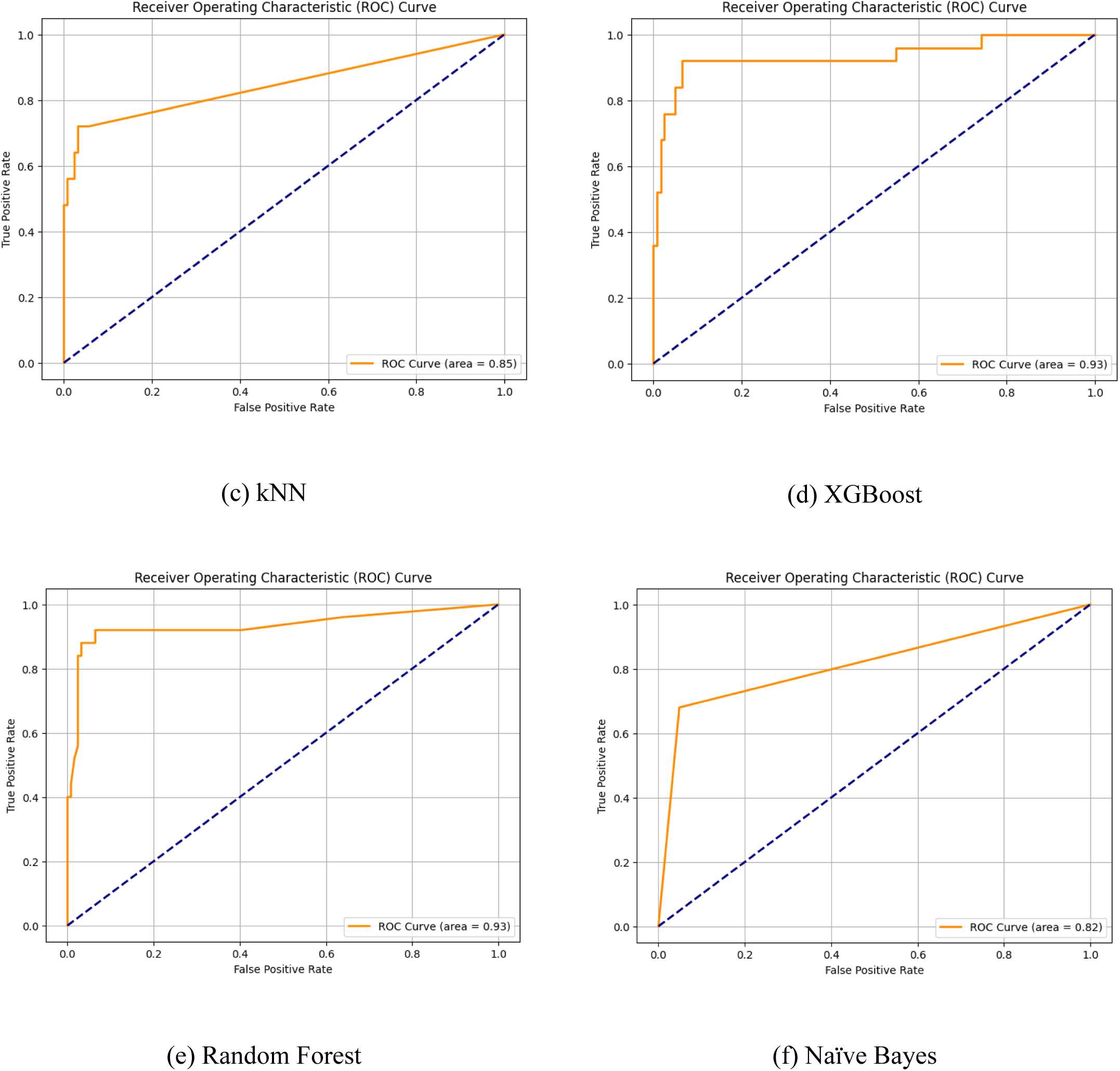
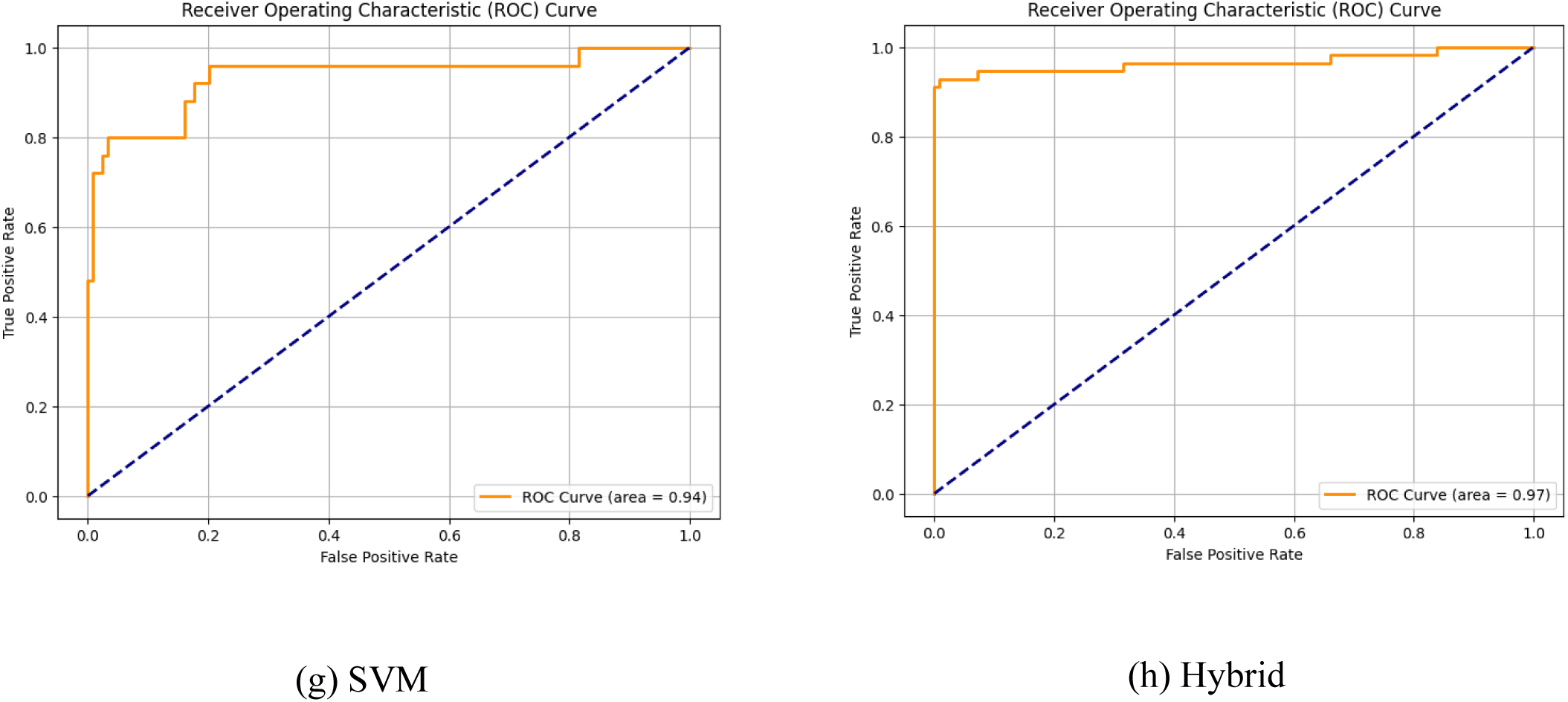
**(a-h). ROC Curve for Different Models**.

The KAN model (AUC = 0.98) and the Hybrid model (AUC = 0.97) have the highest level and show outstanding post-discriminative power. At the second level, CNN (AUC = 0.94), SVM (AUC = 0.94), Random Forest (AUC = 0.93), and XGBoost (AUC = 0.93) models have a good, praiseworthy level of performance. In the last stage, there are KNN (AUC = 0.85) and Naive Bayes (AUC = 0.82). Using the ROC curve alone, the KAN model is superior since its best AUC of 0.98 depicts the strongest ability to classify patients in the widest spectrum of decision thresholds with the utmost accuracy.

#### 4.3.2 Confusion Matrix Analysis

A confusion matrix is an example of a table commonly displayed to describe the results of a classification model on a given test data, where the true values are known. It also enables you to visualize the behavior of an algorithm and can clue you in to the places where the algorithm is faltering. Fig 6 (a-h) shows the different confusion matrices for the 8 different ML models.

**Fig 6.**
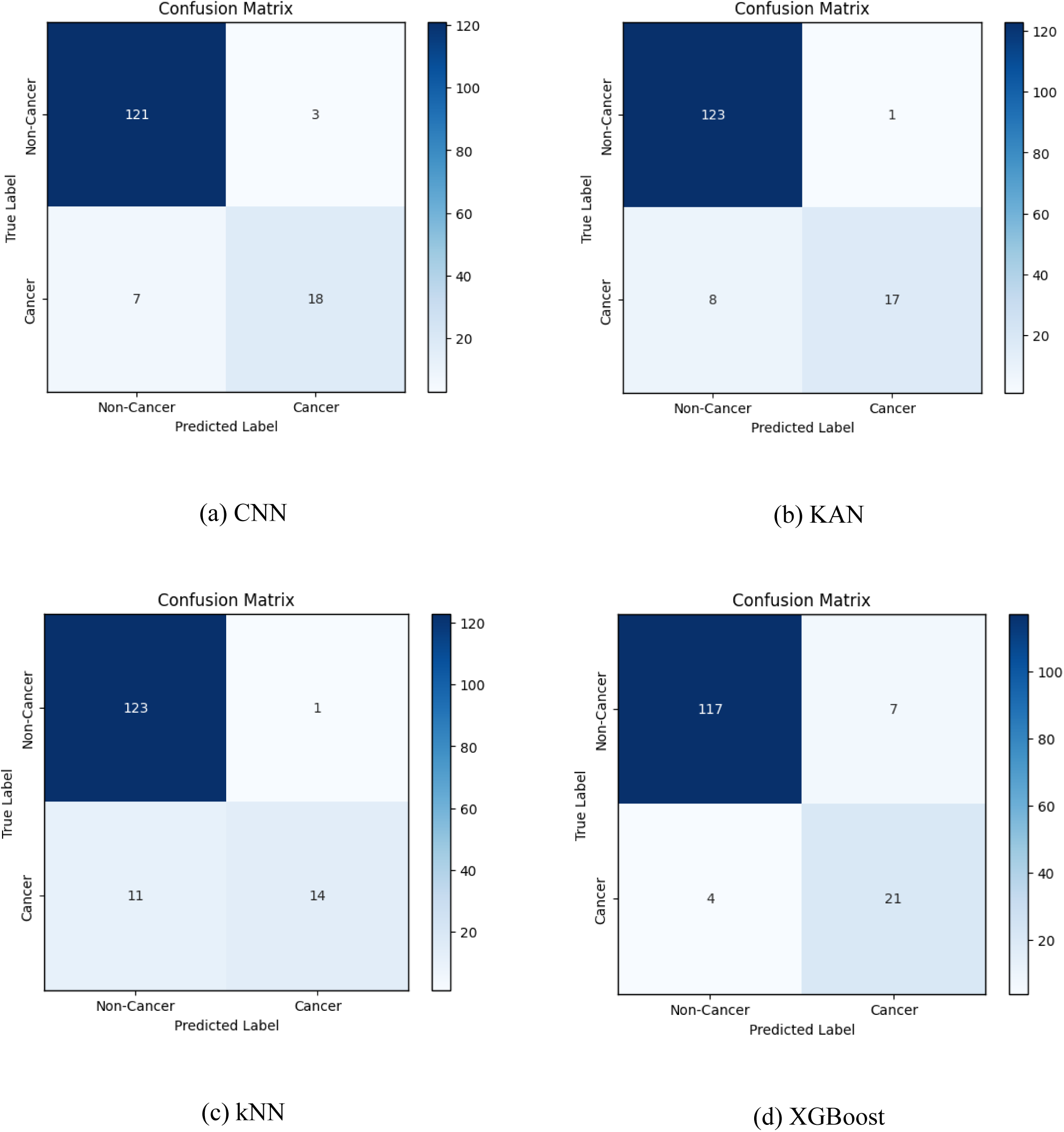
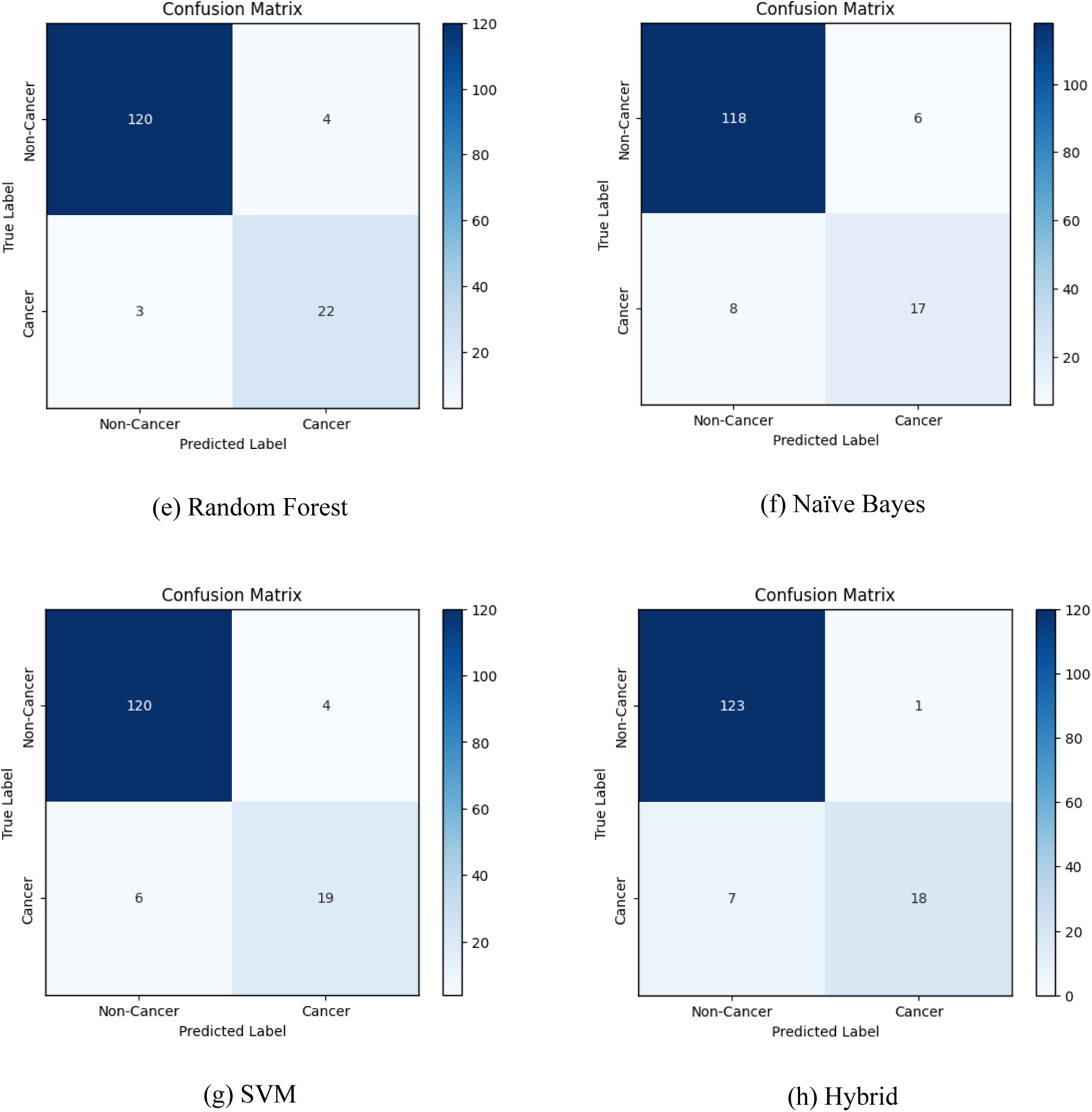
(a-h). Confusion Matrices for Various ML Models.

The model in this situation predicts whether a patient has cancer as follows:

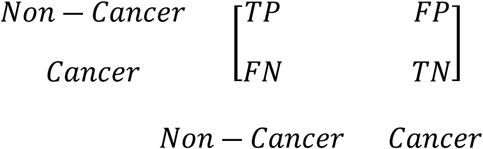

The confusion matrix provides the most direct view of a model’s decision-making, breaking down predictions into True Positives (TP), True Negatives (TN), False Positives (FP), and False Negatives (FN). The values of these are shown in Table 8.

**Table 8.** Confusion Matrix Breakdown for All Evaluated Models.

| Model | True Positive<br>(TP) | True Negative<br>(TN) | False Positive (FP) | False Negative (FN) |
| --- | --- | --- | --- | --- |
| Random Forest | 120 | 22 | 4 | **3** |
| XGBoost | 117 | 21 | 7 | 4 |
| Hybrid | 123 | 18 | **1** | 7 |
| CNN | 121 | 18 | 3 | 7 |
| SVM | 120 | 19 | 4 | 6 |
| KAN | 123 | 17 | **1** | 8 |
| Naïve Bayes | 118 | 17 | 6 | 8 |
| KNN | 123 | 14 | **1** | 11 |

The False Negative (FN) is the most dangerous mistake from a clinical point of view. Here, in such an important feature, the better performance of the Random Forest model is observed, as the false negatives were just 3. However, on the other hand, upon reducing False Positives (FP), the KAN, Hybrid, and kNN models were about 100 percent ahead of the rest, which were not able to identify just one non-cancer case as cancerous, though.

### 4.4 Analysis of Results

The overall analysis shows that one particular model will not perform better in all the criteria than the other unconditionally; the best model depends on the actual and particular aims of a clinical implementation. Nevertheless, a close examination of the findings indicates that in terms of strict statistics and predictive capacity, the Hybrid and KAN models demonstrate the obvious leadership of the introduced designs.

There are several fronts on which the Hybrid and KAN models prove to be superior. The hybrid model has claimed the supremacy owing to its leading Accuracy (0.9667) and F1 Score (0.9444). The most direct measure of performance is the highest accuracy, which ensures it is correct most of the time. Its best F1 Score is even more argumentative, since the latter one admits harmonic mean of the precision and recall demonstrating the high competence of the model to preserve a balanced condition both between the model identifying the true positive cases (high recall) and between the cases when the model predicts positive outcome and the due to it positive judgment (high precision). Such a combination is important in a clinical environment to ensure that a model is sensitive and reliable.

The excellence of the KAN model can be proved in another important group of metrics, which are various. It scored the highest ROC AUC value (0.9760), proving that it has the best inherent ability to differentiate between malignant and benign tissue at all possible diagnostic cut-off points. This serves well as a strong sign of its discriminative power. Moreover, its Log loss score of 0.1421 is the best in class, which means that its probability predictions are calibrated and most assured. This implies that a clinician has more confidence in their probability scores. Its False positive rate is almost perfect, with only one error, and this increases its confidence. This renders it highly sensitive in eliminating cancer, and this eliminates the procedures required later that are unnecessary and stressful.

Notwithstanding the statistical excellence of both models under discussion, the Hybrid and KAN models are not deprived of notable drawbacks that might affect the practical implementation of these models.

#### Limitations of the Hybrid Model

The main limitation of the Hybrid model is that it requires a colossal training cost due to computation. It has a training time of 1009.10 seconds, which makes it one of the most resource-consuming models to develop and retrain. This may be a serious obstacle in research fields or a clinical context, where it would be necessary to iterate on the models quite quickly. Moreover, though its error profile is good, its False Negative count of 6 is twofold the Random Forest model, meaning that it is slightly risky of missing a cancer statistic.

#### Limitations of the KAN Model

The greatest disadvantage of the KAN model is that the model is extremely slow in making its inferences. That is more than 7000 times slower than the Hybrid model and more than 43000 times slower than the Random Forest model, with a time of 0.323 seconds per sample. That would make it inappropriate when considering any actual time - or high-volume screening application. Also, it has a high False Negative rate of 8 as a major trade-off of attaining the lowest FP rate, which has been revealed to be one of the most terrible false negative rates of the highly proficient models. This is a clear safety issue in terms of patients since there is a great risk that the model will not detect a real malignancy relative to some other options.

Conclusively, it should be noted that although the Hybrid and KAN models are practically aloof in their superiority in ability to predict raw predictive power and statistical rigorousness, their drawbacks are substantial in practice. A critical angle is introduced with the analysis of the confusion matrix: Although the Random Forest model had the lowest headline accuracy and AUC, it happened to have the least amount of false negatives. When applying to cancer screening, when the inability to identify a malignancy has the gravest implications, such a low FN rate of the Random Forest model transforms this approach into a strong candidate for implementation. A further practical advantage is that it is extremely efficient. After all, the decision is about the trade-off: the proven statistical superiority of the Hybrid and KAN models on the one hand and the safety profile, yet practical efficiency of the Random Forest model, on the other.

## Chapter 5: Discussion and Recommendations

A thorough investigation was performed in this study that demonstrated a crucial juncture between raw statistical outcomes and actual clinical usability of eight machine learning algorithms in breast cancer detection. The outcome of our experiment clearly shows that even though more powerful and elaborate methods of architecture, such as Hybrid and Kolmogorov-Arnold Network (KAN) models perform significantly better on the most common statistical measures, the Random Forest model offers a more promising and secure framework over which it can find implementation in clinical practice because of its nature of errors. This discussion places these central results in the context of the existing literature, discusses some of the limitations of our best-fitting models, and orients our work to the most urgent knowledge gaps in applied computational medical imaging.

Our Hybrid and KAN models also have a much better performance than the alternative, which is also very consistent with the current prevailing trend in the literature that is seeing a greater attention towards complex, integrated architectures, in place of standalone classifiers. The superior accuracy (0.9667) and F1-score (0.9444) of the Hybrid model highlight the immensely effective synergistic design, most likely, the combined functions of feature extraction (by a deep CNN) and sound decision-making (by an advanced classifier). The methodology has been corroborated in several studies in the recent past, which have been able to use CNNs such as DenseNet121, ResNet50, and VGG16 and seamlessly integrate them with classifiers, which include edRVFL, SVMs, and others, and record state-of-the-art performance [9,13,24]. These results confirm that these models learn and represent the detailed, hierarchical restlessness of mammogram data well, resulting in both very accurate and well-balanced classification performance.

Likewise, it can be noted that the outstanding results of the KAN model (especially its top ROC AUC value (0.9760) and Log Loss (0.1421) show significant potential of innovative neural network paradigms. With the recent advent of KANs, and the U-KAN in particular as the KAN used in their segmentation, much promise has been seen in regards to surpassing the traditional architectures through their increased capabilities to approximate functions better, and provide increased accessibility to interpretation [11]. The findings in our study are good indicators that this is also applicable to high-stakes classification tasks. This high level of accuracy in distinguishing between malignant and benign cases seen in the KAN model in all decision thresholds implies that the KAN model is capturing a deeper insight into the underlying data distribution versus the other models.

Yet, a fatal inconsistency was also established by our analysis as the crux of the scope to adapt AI research to the clinic. Although the Hybrid and KAN models outperformed in terms of statistics, the FN of the Random Forest model was the least (FN=3). A false negative in medical diagnostics would be the failure to diagnose a cancer, the most grave and debilitating type of fallibility. The clinical imperative favors maximum sensitivity (the power to positively identify patients with the disease) at the expense of reduced but only slightly reduced specificity (the power to correctly identify patients without the disease). Thus, compared to the OCR, the Random Forest has a better safety record, which qualifies it as a much more practical and morally acceptable option when it comes to a primary screening method. This result is in line with the general debate in the literature on the pressing need to depart from simple accuracy measures and assess models based on their clinical usefulness in real-life scenarios and their potential risks [14,21].

### 5.1 Limitations

The models with the best performance in our experiment also have considerable limitations; this might be expected, given that the best scores are achieved despite the interesting data on the statistical abilities gap in the research.

Our major constraint in the Hybrid model is that training is computationally demanding and time-consuming (1009.10 sec.). This artifact is reported to be a problem encountered with complex deep learning frameworks with regard to studies that have involved GANs and other deep architectures [8]. The resulting resource requirement is a non-trivial pain point in the clinic or research environment; it may completely block the way to quickly iterate on improvements to the model, to retrain on new data, and to tune hyperparameters widely. This renders the model less dynamic, weak in operating economies, and expensive to upkeep.

Although the KAN model is innovative and strong, it possesses two essential shortcomings thus far, which block its clinical application. First, it took many orders of magnitude longer to make inferences than any other model. This is not a small efficiency issue but could also be an ineluctable impediment to any practical clinical deployment, including performing analysis in real-time, perhaps of a patient visit. Such difficulty is typical of brand-new and intricate models that have not yet been upgraded to speed [17,25]. Second, the high false positive rate (1-specificity) was achieved at the expense of a high false negative rate (FN=8). This is inherent in classifier performance where precision versus sensitivity causes a trade-off, but in this instance, there is a strong tendency to lean towards the decisions that are comfortable, but not towards patient safety, which is an essential that must be resolved before this or any model could even be considered for clinical application.

In addition to this, our research possesses the same shortcomings as most of the studies in the field. We work with one univariate and limited data set. As already stressed in many studies, external validation on larger, multi-institutional and more diverse datasets is critical and sustained through the future to build confidence in model generalizability and robustness [6,7,9,16,23]. In the absence of such validation, it is highly likely that the performance of a model is merely due to overfitting on the idiosyncrasies of a training data set, and any test of it on images belonging to other equipment manufacturers or patient cohorts may prove inadequate. The inability to integrate other data modalities, namely patient clinical history, genomic data, or pathology reports, is also a strong missed opportunity for better performance, which [6] and [16] also illustrate.

### 5.3 Recommendations for Future Research

Based on our findings and the gaps discovered in the literature, we arrived at the specific and actionable future research propositions as follows:

#### Optimization and Hybridization of Novel Architectures

The future of this work should focus more on optimization of the KAN architecture to come up with a real time drastically reduced inference time. To achieve this step, it may be worth investigating methods of model pruning, quantization, or knowledge distillation in order to make KANs useful in a clinical setting. Also, the current Hybrid model and the U-KAN architecture [11] that we have witnessed success strongly points out that integrating KANs with any popular CNN may be a productive direction. An integration of CNNs (using CNN-KANs) would, in theory, attempt to take advantage of the established feature extraction advantages of CNNs, together with the higher discriminative capabilities and interpretability supplied by KANs.

#### Emphasis on ‘Clinically-Aware Ensembles’

The next step should be to work on sophisticated ensemble techniques that have been specifically tailored to avoid prevalent catastrophic types of error. As an example, we could make an ensemble, which places more emphasis on the models with low F-Negative (as in our case, Random Forest), but when it comes to the intermediate cases, it uses the models with low F-Positive (KAN in our case) to settle the dispute.

#### Priority of Data Diversity and Generalizability

The much-needed emphasis on model generalizability presents an opportunity to be filled in by the research community with utmost priority. Future research should no longer be based on single-institutional data; rather, efforts should be shifted to acquiring and using large-scale data, multi-institutional data, and multi-vendor data. This echoes an action that is frequently reflected in the literature [8,14] and is necessary to create AI tools that are robust, fair, and effective in all patient groups, not only those ones, which are included in the original training cohort.

#### Intensive Multi-Modal Data Integration

To reach yet another step up in the level of predictive accuracy, in the future, the models should go past image-based analysis and strive towards integrating imaging data and other rich information sources. A genuinely holistic model would include not only structured data (clinical data, lab results) and unstructured data (pathology reports), but even genomic or proteomic data, as is implied by [6] and [22]. In such an approach, individualized, context-sensitive, and more inductive diagnostic tools can be developed.

#### Through Human-Centric Explainability (XAI)

Human-Centric Explainability (XAI), as AI models grow more complex by the day. The black box nature of the model poses a serious challenge to clinical implementation. A model should not only be accurate, but it should also be able to have confidence in it and be able to know how it arrived at its conclusions. Future efforts should surpass work producing plain saliency maps [6,21], and more sophisticated forms of XAI need to be generated. These may involve ways of offering textual clarifications, showing precedent cases, or measuring the uncertainty of the model of a particular prediction, therefore, establishing credibility and easing the responsible application.

In the future, these models could be tested on different, multi-institutional data sets to confirm the extent to which they are effective in various populations and under different imaging situations in the future. Inference performance of lightweight models such as the Kolmogorov-Arnold Network may be improved to increase general applicability to real-time diagnostics by means of algorithmic and hardware optimizations. Investigating multi-class categorization to differentiate the subtypes or stages of cancer may give more specific results, which may result in improved clinical decision-making. Adding interpretability mechanisms, like attention maps or feature importance analysis, might be of value in getting clinicians more confident in model outputs, and thereby adopted into medical practice. Moreover, identifying a way to minimize the resources required by the high-performing model, e.g., using model compression methods or edge computing type, might open the way to utilizing these models in the low-resource environments where they are most needed, namely, screening breast cancer and meeting the global needs. This integration of machine learning with clinical workflows will also be promoted by these initiatives, and it will eventually lead to better early detection and patient care as well.

The comparative analysis offered in this work critically compares two distinct performance areas (statistical performance and clinical safety) and relative efficiency in the strategic field of breast cancer diagnosis due to eight machine learning models, which are accompanied by the strengths and weaknesses, and nuances of their usage. We found out that advanced and novel architectures are statistically better, and the Hybrid and KAN models have a state-of-the-art predictive strength. We, however, also emphasize the utmost significance of granular error analysis, which can also be seen as a rather strong argument in favor of the computationally efficient and extremely sensitive Random Forest model being on the safer and more appropriate side of the spectrum of a properly functioning primary clinical screening tool. This work ultimately reinstates the fact that translation of artificial intelligence applications between the laboratory and clinic is a complex process that needs a circumferential analysis tool-a framework that is keen on weighing both technical perfection and practicality of applications in terms of computation resources and, first and foremost, the inescapable ethical direction of patient security.

## Data Availability

The data used maybe found in the reference section of the manuscript.

https://data.mendeley.com/datasets/fvjhtskg93/1

